# Distinct smell and taste disorder phenotype of post-acute COVID-19 sequelae

**DOI:** 10.1101/2022.06.02.22275932

**Authors:** Verena Rass, Piotr Tymoszuk, Sabina Sahanic, Beatrice Heim, Dietmar Ausserhofer, Anna Lindner, Mario Kofler, Philipp Mahlknecht, Anna Boehm, Katharina Hüfner, Alex Pizzini, Thomas Sonnweber, Katharina Kurz, Bernhard Pfeifer, Stefan Kiechl, Marina Peball, Philipp Kindl, Lauma Putnina, Elena Fava, Atbin Djamshidian, Andreas Huber, Christian J. Wiedermann, Barbara Sperner-Unterweger, Ewald Wöll, Ronny Beer, Alois Josef Schiefecker, Rosa Bellmann-Weiler, Herbert Bachler, Ivan Tancevski, Bettina Pfausler, Giuliano Piccoliori, Klaus Seppi, Günter Weiss, Judith Löffler-Ragg, Raimund Helbok

**Affiliations:** Department of Neurology, Medical University of Innsbruck, Innsbruck, Austria; Data Analytics As a Service Tirol, Innsbruck, Austria; Department of Internal Medicine II, Medical University of Innsbruck, Innsbruck Austria; Institute of General Practice and Public Health, Claudiana College of Health Professions, Bolzano, Italy; Department of Psychiatry, Psychotherapy, Psychosomatics and Medical Psychology, University Hospital for Psychiatry II, Medical University of Innsbruck, Innsbruck, Austria; Tyrolean Federal Institute for Integrated Care, Innsbruck, Austria; Division for Health Networking and Telehealth, Biomedical Informatics and Mechatronics, UMIT, Hall in Tyrol, Austria; Department of Internal Medicine, St. Vinzenz Hospital, Zams, Austria; Institute of General Medicine, Medical University of Innsbruck, Innsbruck, Austria

**Keywords:** olfactory dysfunction, COVID-19, long COVID, mental health, quality of life

## Abstract

**Background:** Olfactory dysfunction (OD) often accompanies acute coronavirus disease 2019 (COVID-19) and its sequelae. Herein, we investigated OD during COVID-19 recovery in the context of other symptoms, quality of life, physical and mental health.

**Methods:** Symptom recovery patterns were analyzed in a bi-national, ambulatory COVID-19 survey (n = 906, ≥ 90 days follow-up) and a multi-center observational cross-sectional cohort of ambulatory and hospitalized individuals (n = 108, 360 days follow-up) with multi-dimensional scaling, association rule mining and partitioning around medoids clustering.

**Results:** Both in the ambulatory collective (72%, n = 655/906) and the cross-sectional ambulatory and hospitalized cohort (41%, n = 44/108) self-reported OD was frequent during acute COVID-19, displayed a slow recovery pace (ambulatory: 28 days, cross-sectional: 90 days median recovery time) and commonly co-occurred with taste disorders. In the ambulatory collective, a predominantly young, female, comorbidity-free group of convalescents with persistent OD and taste disorder (>90 days) was identified. This post-acute smell and taste disorder phenotype was characterized by a low frequency of other leading post-acute symptoms including fatigue, respiratory and neurocognitive complaints. Despite a protracted smell and taste dysfunction, this subset had high ratings of physical performance, mental health, and quality of life.

**Conclusion:** Our results underline the clinical heterogeneity of post-acute COVID-19 sequelae calling for tailored management strategies. The persistent smell and taste disorder phenotype may represent a distinct COVID-19 recovery pathway characterized by a good recovery of other COVID-19 related symptoms.

**Study registration:** ClinicalTrials.gov: NCT04661462 (ambulatory collective), NCT04416100 (cross-sectional cohort).

## Introduction

Coronavirus disease 2019 (COVID-19) manifests with various respiratory, neurological, neurocognitive and cardiopulmonary symptoms (1–3). A considerable number of COVID-19 patients suffers from persistent symptoms with implications on quality of life and mental health (3– 7). All post-COVID-19 symptoms over 28 days are subsumed under the patient-coined term ‘long COVID’ (8). Furthermore persistent symptoms present for > 12 weeks were coarsely classified as PASC (post-acute sequelae of COVID-19) (9,10). However, these definitions do not address the character of persistent symptoms or their burden.

Olfactory dysfunction (OD) is a frequent symptom of acute COVID-19 (11), in particular for the wild type, alpha and delta variants of the SARS-CoV-2 virus (12,13). Such COVID-19-related OD may result from injury of upper respiratory epithelial cells or neurons of the olfactory mucosa, olfactory bulb, primary olfactory cortex or secondary projection areas (14,15). OD during acute COVID-19 is estimated to affect up to 48% patients (16). Although literature suggests resolution of OD within 2-3 weeks in most patients (17), OD may persist up to 6 months in 5-11% of patients (11,18–20). Consequently, it represents an important post-acute sequelae, with a disabling character for certain patients due to its effect on quality of life, daily functioning and professional activity (20–23). Characterization of COVID-19 recovery patterns is crucial to identify patients at risk of persistent OD who may profit from targeted therapy such as olfactory training.

Herein, we investigated clinical and psychosocial recovery in ambulatory and hospitalized COVID-19 patients with a particular focus on OD. We therefore re-analyzed our previously published bi-national survey of non-hospitalized COVID-19 patients (Health after COVID-19 in Tyrol, HACT) (1,6) and a multi-center observational study including both ambulatory and hospitalized patients (CovILD) (3,5,24) recruited during outbreaks of wild type and alpha variant of the SARS-CoV-2 pathogen with association mining and clustering algorithms.

## Methods

### Study design and approval

The HACT online survey (ClinicalTrials.gov: NCT04661462) encompassed two independently recruited cohorts of ambulatory COVID-19 convalescents in the provinces Tyrol (Austria, AT) and South Tyrol (Italy, IT). Adult residents of the study regions with a laboratory-confirmed SARS-CoV-2 infection were included in the study. Hospitalized COVID-19 patients were excluded (1,6). The participants completed the survey between 30^th^ September 2020 and 5^th^ July 2021, i. e. during the predominance of the wild-type and alpha variant of SARS-CoV-2 in the study regions, as discussed before (1,25). The HACT study subset with symptomatic COVID-19 and ≥90 days between diagnosis and survey completion was included in the analysis (AT: n = 479, IT: n = 427, **Figure 1**).

**Figure 1.**
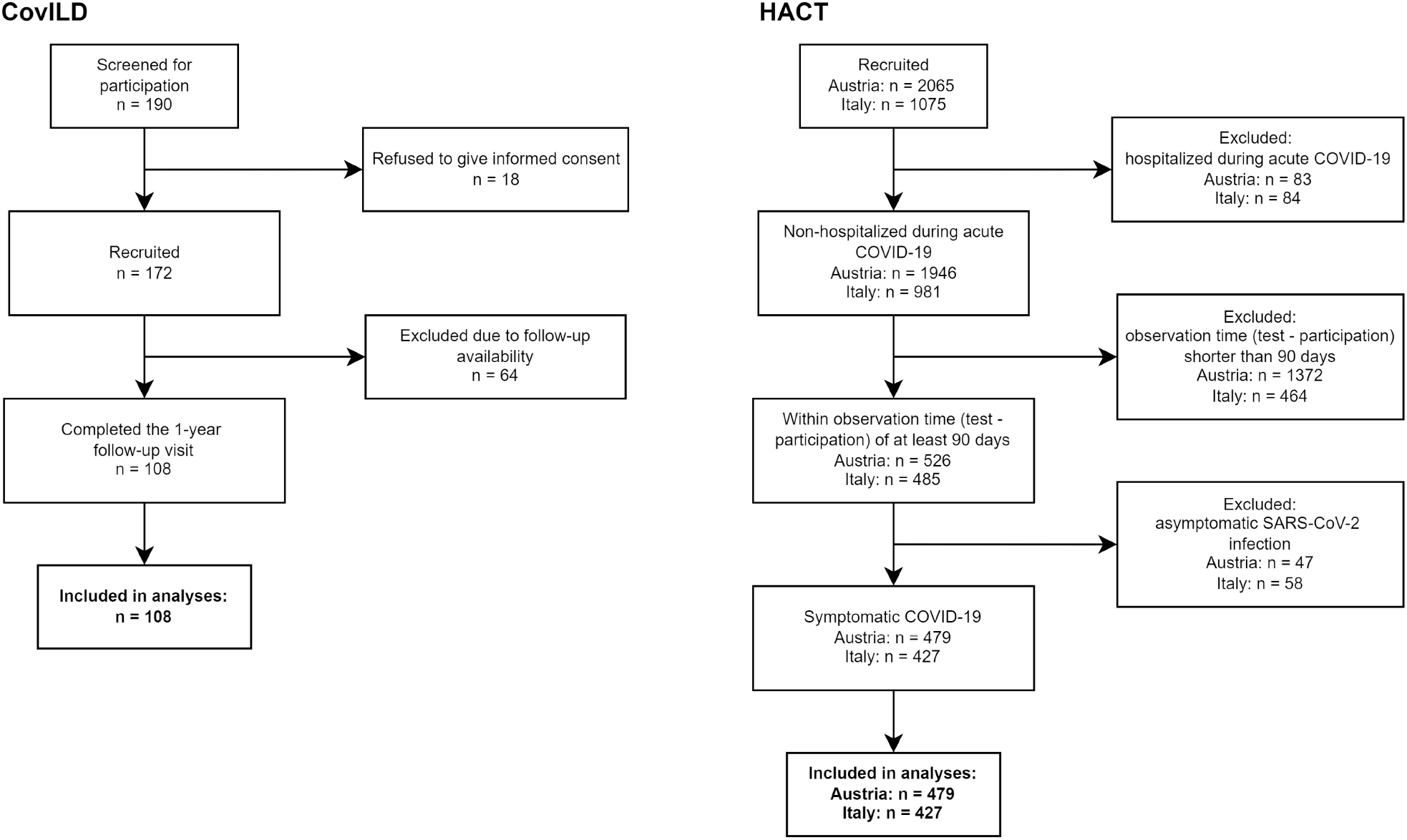
Flow diagram of the analysis inclusion process for the CovILD and HACT studies..

The CovILD longitudinal observational cohort (NCT04416100) included both COVID-19 outpatients and inpatients recruited at the Department of Internal Medicine II at the Medical University of Innsbruck, St. Vinzenz Hospital in Zams and the acute rehabilitation facility in Muenster (all in Austria) between March and June 2020 (3,5,24). To this date, no variants of SARS-CoV-2 were reported (25). The study data for acute COVID-19 and the 60-, 100-, 180-and 360-day follow-ups for participants who had completed all follow-ups were analyzed (n = 108, **Figure 1**).

The studies were conducted in accordance with the Declaration of Helsinki and European Data Policy. All participants gave digitally signed or written informed consent prior to enrollment. The study protocols were reviewed and approved by the institutional review boards of the Medical University of Innsbruck (HACT AT, approval number: 1257/2020, CovILD: 1103/2020) and of the Autonomous Province of Bolzano – South Tyrol (HACT IT, approval number: 0150701).

### Procedures and study variables

In the HACT online survey, participants assigned 42 symptoms to pre-defined duration classes (absent: re-coded as 0 days, 1 – 3 days: 3 days, up to 1 week: 7 days, up to 2 weeks: 14 days, up to 4 weeks: 28 days, up to 3 months: 90 days, up to 6 months: 90 days, over 6 months: 90 days). The symptoms were classified as acute (present during 14 days after clinical onset), long COVID (≥28 days) and PASC (post-acute sequelae of COVID-19, ≥90 days) manifestations (1,6,9). Self-perceived complete recovery, need for rehabilitation and new drugs since COVID-19 were surveyed as single yes/no items. Percentage of physical performance loss following COVID-19 was rated with a 0 – 100% scale (1,6). Quality of life impairment (4-item likert scale), overall mental health impairment (4-item likert scale), anxiety and depression (Patient Health Questionnaire, PHQ-4) and psychosocial stress (7 item PHQ stress module) at the time of survey completion were assessed as described before (1,6,26,27).

In the CovILD study, the presence of 8 symptoms (reduced physical performance, hyposmia/anosmia, dyspnea, sleep problems, cough, fever, night sweating, gastrointestinal symptoms) were prospectively recorded at each of 60-, 100-, 180-and 360-day post COVID-19 follow-up. Acute COVID-19 symptoms were assessed retrospectively (3,5). Objective olfactory function at the 100-and 360-day follow-up was assessed with the 16-item Sniffin’ Sticks Identification test (Burghart Medizintechnik, Germany). Objective hyposmia was defined at a cutoff of <13 points as per manufacturer criteria (24,28).

Study variables are listed in **Supplementary Table S1** (HACT) and **S2** (CovILD).

### Statistical analysis

Data transformation, statistical analysis and result visualization was conducted with R, version 4.0.5 (29–31). Statistical significance for differences in numeric variables was investigated by Mann-Whitney U test with Wilcoxon r effect size statistic of Kruskal-Wallis test with η^2^ effect size statistic, as appropriate (package *rstatix*) (32). Differences in categorical variable frequency were assessed by χ^2^ test with Cramer V effect size statistic (*rstatix*) (32). Concordance between subjective and objective hyposmia was measured with Cohen’s κ statistic (package *vcd*) (33). Two-dimensional multi-dimensional scaling (MDS) was performed with simple matching distance matrices for binary symptom data (0: absent, 1: present, package *scrime* and base R *cmdscale()* function) (34,35). Apriori association rule analysis for frequent symptom combinations was conducted with *arules* package (36,37). Clustering of the training HACT AT cohort by symptom-specific recovery times was done with the PAM algorithm (partitioning around medoids, Euclidean distance, packages *philentropy* and *cluster*) (38,39). Assignment of the test IT cohort participants to the AT cohort-defined clusters was accomplished with the 5-nearest neighbors classification algorithm (40). P values were corrected for multiple testing with Benjamini-Hochberg method (41). For details of the statistical analysis, see **Supplementary Methods**.

## Results

### Study population

Out of 3140 HACT study respondents (1,6), 906 non-hospitalized individuals with symptomatic COVID-19 surveyed ≥90 days after diagnosis were included in the analysis. Those individuals were grouped in two independently recruited national Austria and Italy cohorts (AT: n = 479 and IT n = 427) (**Figure 1**). The median follow-up time ranged from 140 (IT) to 180 days (AT). The cohorts consisted primarily of working-age individuals (AT: median 43 [IQR: 32 – 53]; IT: median 45 [34 – 54] years) and females were over-represented (AT: 67%, IT: 70%). Almost half of the participants had at least one comorbidity (AT: 49%, IT: 43%), with hay fever/allergy, arterial hypertension and obesity (body mass index >30 kg/m^2^) being the most frequent conditions (**Table 1**).

**Table 1:**
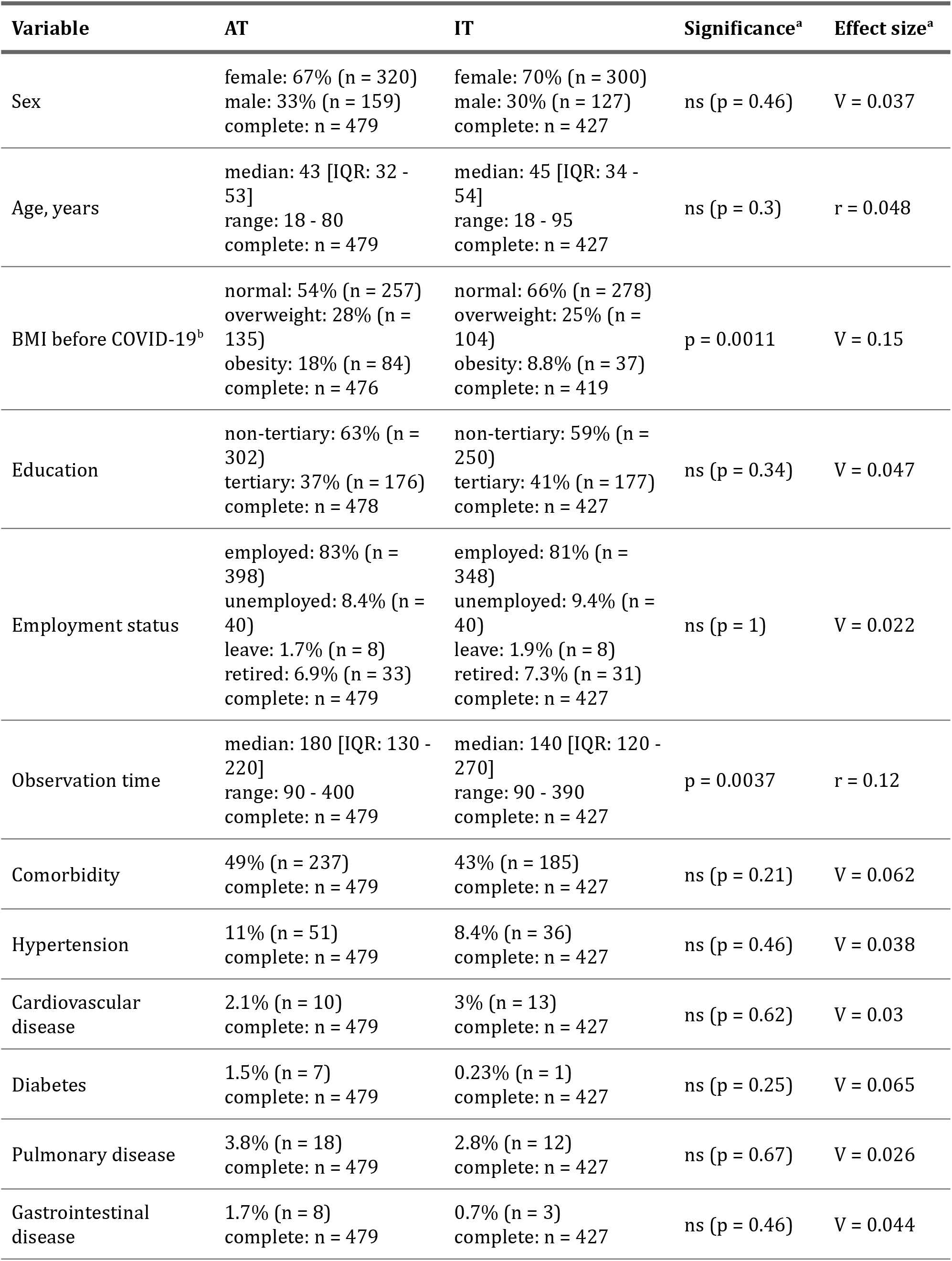
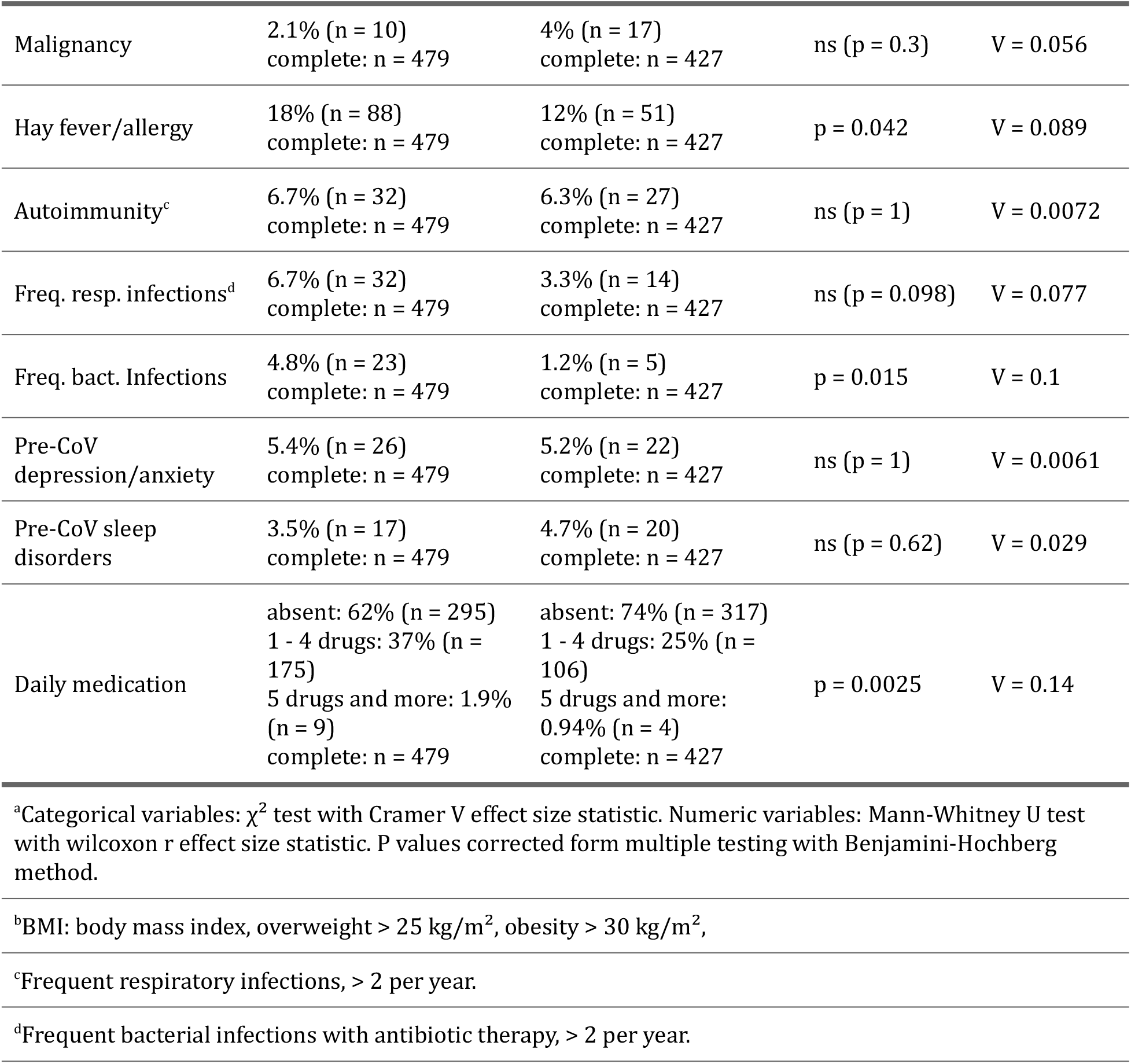
Baseline characteristic of the HACT study Austria (AT) and Italy (IT) cohorts.

| Variable | AT | IT | Significance<sup>a</sup> | Effect size<sup>a</sup> |
| --- | --- | --- | --- | --- |
| Sex | female: 67% (n = 320)<br>male: 33% (n = 159)<br>complete: n = 479 | female: 70% (n = 300)<br>male: 30% (n = 127)<br>complete: n = 427 | ns (p = 0.46) | V = 0.037 |
| Age, years | median: 43 [IQR: 32 - 53]<br>range: 18 - 80<br>complete: n = 479 | median: 45 [IQR: 34 - 54]<br>range: 18 - 95<br>complete: n = 427 | ns (p = 0.3) | r = 0.048 |
| BMI before COVID-19<sup>b</sup> | normal: 54% (n = 257)<br>overweight: 28% (n = 135)<br>obesity: 18% (n = 84)<br>complete: n = 476 | normal: 66% (n = 278)<br>overweight: 25% (n = 104)<br>obesity: 8.8% (n = 37)<br>complete: n = 419 | p = 0.0011 | V = 0.15 |
| Education | non-tertiary: 63% (n = 302)<br>tertiary: 37% (n = 176)<br>complete: n = 478 | non-tertiary: 59% (n = 250)<br>tertiary: 41% (n = 177)<br>complete: n = 427 | ns (p = 0.34) | V = 0.047 |
| Employment status | employed: 83% (n = 398)<br>unemployed: 8.4% (n = 40)<br>leave: 1.7% (n = 8)<br>retired: 6.9% (n = 33)<br>complete: n = 479 | employed: 81% (n = 348)<br>unemployed: 9.4% (n = 40)<br>leave: 1.9% (n = 8)<br>retired: 7.3% (n = 31)<br>complete: n = 427 | ns (p = 1) | V = 0.022 |
| Observation time | median: 180 [IQR: 130 - 220]<br>range: 90 - 400<br>complete: n = 479 | median: 140 [IQR: 120 - 270]<br>range: 90 - 390<br>complete: n = 427 | p = 0.0037 | r = 0.12 |
| Comorbidity | 49% (n = 237)<br>complete: n = 479 | 43% (n = 185)<br>complete: n = 427 | ns (p = 0.21) | V = 0.062 |
| Hypertension | 11% (n = 51)<br>complete: n = 479 | 8.4% (n = 36)<br>complete: n = 427 | ns (p = 0.46) | V = 0.038 |
| Cardiovascular disease | 2.1% (n = 10)<br>complete: n = 479 | 3% (n = 13)<br>complete: n = 427 | ns (p = 0.62) | V = 0.03 |
| Diabetes | 1.5% (n = 7)<br>complete: n = 479 | 0.23% (n = 1)<br>complete: n = 427 | ns (p = 0.25) | V = 0.065 |
| Pulmonary disease | 3.8% (n = 18)<br>complete: n = 479 | 2.8% (n = 12)<br>complete: n = 427 | ns (p = 0.67) | V = 0.026 |
| Gastrointestinal disease | 1.7% (n = 8)<br>complete: n = 479 | 0.7% (n = 3)<br>complete: n = 427 | ns (p = 0.46) | V = 0.044 |
| Malignancy | 2.1% (n = 10)<br>complete: n = 479 | 4% (n = 17)<br>complete: n = 427 | ns (p = 0.3) | V = 0.056 |
| Hay fever/allergy | 18% (n = 88)<br>complete: n = 479 | 12% (n = 51)<br>complete: n = 427 | p = 0.042 | V = 0.089 |
| Autoimmunity<sup>c</sup> | 6.7% (n = 32)<br>complete: n = 479 | 6.3% (n = 27)<br>complete: n = 427 | ns (p = 1) | V = 0.0072 |
| Freq. resp. infections<sup>d</sup> | 6.7% (n = 32)<br>complete: n = 479 | 3.3% (n = 14)<br>complete: n = 427 | ns (p = 0.098) | V = 0.077 |
| Freq. bact. Infections | 4.8% (n = 23)<br>complete: n = 479 | 1.2% (n = 5)<br>complete: n = 427 | p = 0.015 | V = 0.1 |
| Pre-CoV depression/anxiety | 5.4% (n = 26)<br>complete: n = 479 | 5.2% (n = 22)<br>complete: n = 427 | ns (p = 1) | V = 0.0061 |
| Pre-CoV sleep disorders | 3.5% (n = 17)<br>complete: n = 479 | 4.7% (n = 20)<br>complete: n = 427 | ns (p = 0.62) | V = 0.029 |
| Daily medication | absent: 62% (n = 295)<br>1 - 4 drugs: 37% (n = 175)<br>5 drugs and more: 1.9% (n = 9)<br>complete: n = 479 | absent: 74% (n = 317)<br>1 - 4 drugs: 25% (n = 106)<br>5 drugs and more: 0.94% (n = 4)<br>complete: n = 427 | p = 0.0025 | V = 0.14 |

<sup>a</sup>Categorical variables: $\chi^2$ test with Cramer V effect size statistic. Numeric variables: Mann-Whitney U test with wilcoxon r effect size statistic. P values corrected form multiple testing with Benjamini-Hochberg method.
<sup>b</sup>BMI: body mass index, overweight > 25 kg/m<sup>2</sup>, obesity > 30 kg/m<sup>2</sup>,
<sup>c</sup>Frequent respiratory infections, > 2 per year.
<sup>d</sup>Frequent bacterial infections with antibiotic therapy, > 2 per year.

CovILD study participants who underwent all follow-ups up to one year after COVID-19 (n = 108, **Figure 1**) were predominantly male (59%). The median age was 56 (IQR: 49 – 68) years and 75% of the participants had comorbidities such as obesity, cardiovascular or pulmonary disease or type 2 diabetes mellitus. The CovILD study participants were stratified as mild (outpatients, 25%), moderate (inpatients, no mechanical ventilation or ICU, 51%) and severe COVID-19 convalescents (mechanical ventilation or ICU, 24%). The median age and comorbidity rates were significantly higher in moderate or severe COVID-19 survivors compared to mild disease patients (**Table 2**).

**Table 2:** Baseline characteristic of the CovILD study cohort and the CovILD study participants stratified by COVID-19 severity.

| Variable | Entire cohort | Ambulatory CoV subset | Moderate CoV subset | Severe CoV subset | Significance<sup>a</sup> | Effect size<sup>a</sup> |
| --- | --- | --- | --- | --- | --- | --- |
| Sex | female: 41% (n = 44)<br>male: 59% (n = 64)<br>complete: n = 108 | female: 67% (n = 18)<br>male: 33% (n = 9)<br>complete: n = 27 | female: 35% (n = 19)<br>male: 65% (n = 36)<br>complete: n = 55 | female: 27% (n = 7)<br>male: 73% (n = 19)<br>complete: n = 26 | p < 0.001 | V = 0.31 |
| Age, years | median: 56<br>[IQR: 49 - 68]<br>range: 19 - 87<br>complete: n = 108 | median: 47<br>[IQR: 38 - 55]<br>range: 19 - 70<br>complete: n = 27 | median: 62<br>[IQR: 53 - 72]<br>range: 27 - 87<br>complete: n = 55 | median: 56<br>[IQR: 52 - 64]<br>range: 44 - 79<br>complete: n = 26 | p < 0.001 | $\eta^2 = 0.21$ |
| BMI at CoV onset<sup>b</sup> | normal: 39% (n = 42)<br>overweight: 43% (n = 46)<br>obesity: 19% (n = 20)<br>complete: n = 108 | normal: 56% (n = 15)<br>overweight: 33% (n = 9)<br>obesity: 11% (n = 3)<br>complete: n = 27 | normal: 29% (n = 16)<br>overweight: 51% (n = 28)<br>obesity: 20% (n = 11)<br>complete: n = 55 | normal: 42% (n = 11)<br>overweight: 35% (n = 9)<br>obesity: 23% (n = 6)<br>complete: n = 26 | p < 0.001 | V = 0.17 |
| Comorbidity present | 75% (n = 81)<br>complete: n = 108 | 41% (n = 11)<br>complete: n = 27 | 85% (n = 47)<br>complete: n = 55 | 88% (n = 23)<br>complete: n = 26 | p < 0.001 | V = 0.46 |
| Metabolic disease | 42% (n = 45)<br>complete: n = 108 | 19% (n = 5)<br>complete: n = 27 | 49% (n = 27)<br>complete: n = 55 | 50% (n = 13)<br>complete: n = 26 | p < 0.001 | V = 0.27 |
| Hypertension | 27% (n = 29)<br>complete: n = 108 | 7.4% (n = 2)<br>complete: n = 27 | 27% (n = 15)<br>complete: n = 55 | 46% (n = 12)<br>complete: n = 26 | p < 0.001 | V = 0.31 |
| Cardiovascular disease | 40% (n = 43)<br>complete: n = 108 | 7.4% (n = 2)<br>complete: n = 27 | 47% (n = 26)<br>complete: n = 55 | 58% (n = 15)<br>complete: n = 26 | p < 0.001 | V = 0.39 |
| Diabetes | 15% (n = 16)<br>complete: n = 108 | 3.7% (n = 1)<br>complete: n = 27 | 15% (n = 8)<br>complete: n = 55 | 27% (n = 7)<br>complete: n = 26 | p < 0.001 | V = 0.23 |
| Pulmonary disease | 19% (n = 20)<br>complete: n = 108 | 11% (n = 3)<br>complete: n = 27 | 22% (n = 12)<br>complete: n = 55 | 19% (n = 5)<br>complete: n = 26 | p = 0.031 | V = 0.11 |
| Gastrointestinal disease | 13% (n = 14)<br>complete: n = 108 | 0% (n = 0)<br>complete: n = 27 | 20% (n = 11)<br>complete: n = 55 | 12% (n = 3)<br>complete: n = 26 | p < 0.001 | V = 0.24 |
| Malignancy | 9.3% (n = 10)<br>complete: n = 108 | 3.7% (n = 1)<br>complete: n = 27 | 15% (n = 8)<br>complete: n = 55 | 3.8% (n = 1)<br>complete: n = 26 | p < 0.001 | V = 0.19 |
| Immune deficiency | 5.6% (n = 6)<br>complete: n = 108 | 0% (n = 0)<br>complete: n = 27 | 3.6% (n = 2)<br>complete: n = 55 | 15% (n = 4)<br>complete: n = 26 | p < 0.001 | V = 0.25 |

<sup>a</sup>Comparison of ambulatory, moderate and severe COVID-19 individuals. Categorical variables: $\chi^2$ test with Cramer V effect size statistic. Numeric variables: Kruskal-Wallis test with $\eta^2$ effect size statistic. P values corrected form multiple testing with Benjamini-Hochberg method.
<sup>b</sup>BMI: body mass index, overweight > 25 kg/m<sup>2</sup>, obesity > 30 kg/m<sup>2</sup>,

### Longitudinal course of COVID-19 symptom resolution

During acute COVID-19, subjective smell (AT: 70%, IT: 75%) and self-reported taste disorders (AT: 68%, IT: 74%) along with fatigue, tiredness, diminished appetite, joint pain, tachypnea, cough and fever were present in the majority of the HACT study participants. While most upper airway and infection symptoms resolved rapidly, OD and taste deficits resolved substantially slower with median recovery times of 14 (taste) to 28 days (smell disorders) (**Figure 2, Supplementary Figure S1 – S2**). Accordingly, OD (AT: 30%, IT: 27%) and taste disorders (AT: 22%, IT: 21%) were still present in more than one-fifth of the convalescents 90 days after COVID-19 onset. Other symptoms with prolonged recovery times included memory and concentration impairment, tachypnea, tiredness and fatigue (**Figure 2, Supplementary Figure S1 – S2**).

**Figure 2.**
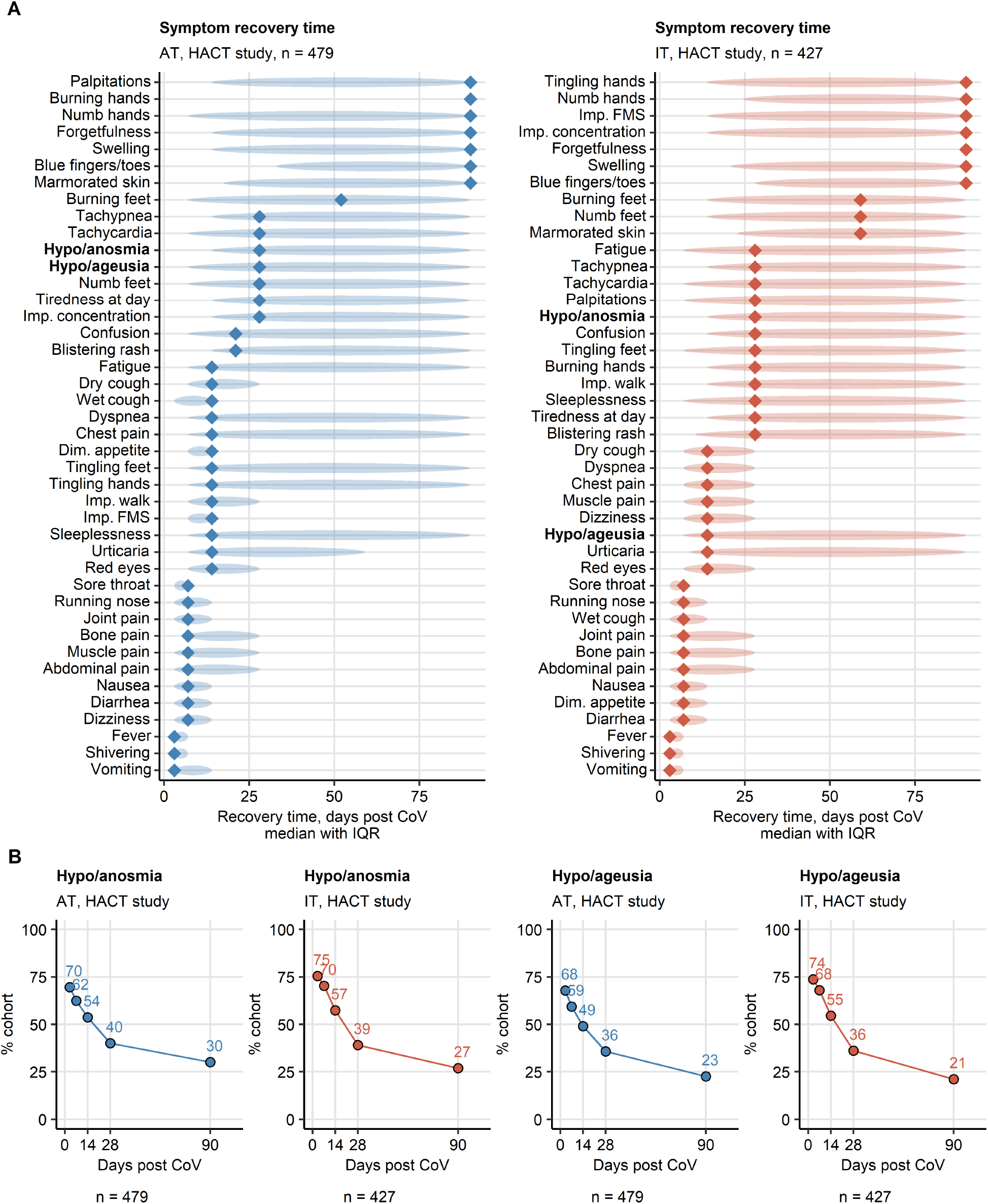
Symptom-specific recovery times in ambulatory COVID-19. Symptom-specific recovery times were calculated for each participants of the HACT study cohorts (Austria: AT, Italy: IT). Imp.: impaired, Dim.: diminished, FMS: fine motor skills. **(A)** Distribution of the recovery times in the individuals with the indicated symptoms present during acute COVID-19. Diamonds represent median recovery times, tinted ellipses code for interquartile ranges. Numbers of complete observations are indicated in the plot captions. **(B)** Percentages of individuals with smell and taste disorders in the AT (Austria) and IT (Italy) HACT study cohorts at particular time points after clinical onset. Numbers of complete observations are indicated under the plots.

Similarly, self-reported OD was a frequent acute COVID-19 symptom in both inpatients and outpatients of the CovILD cohort (mild: 47%, moderate: 33%, severe COVID-19: 53%) (**Supplementary Figure S3**). The median recovery time across all disease severity strata was 90 days. Despite a significant resolution over the first year, OD was characterized by a high chronicity in mild or moderate COVID-19 convalescents (16% at the 360-day follow-up). By contrast, recovery of self-perceived OD was substantially faster in severe COVID-19 resulting in a complete resolution within 6 months in all patients (**Supplementary Figures S3 – S4**).

### Comparison of subjective and objective OD

Objective OD (Sniffin’ stick test < 13 points) (24,28) in the entire CovILD collective was higher both at the 100-(45%, n = 95 individuals tested) and 360-day (54%, n = 63 tested) follow-up compared to self-reported OD (100-days follow-up: 18%, 360-day: 9.5%). This was also true for the COVID-19 severity strata. In this line, a fair significant association of objective and subjective OD was observed solely in the moderate COVID-19 convalescents (100 day follow-up: κ = 0.32 [95%CI: 0.091 – 0.56], 360 day: κ = 0.29 [0.057 – 0.52]). The most striking difference was found in severe COVID-19 patients at the 360-day follow-up where 71% without the perception of subjective OD were diagnosed with objective OD (κ = 0) (**Supplementary Figures S5 – S6**).

### Subjective OD and taste disorders as distinct post-acute sequelae of COVID-19 in the HACT study

As discerned by mapping of symptom-symptom simple matching distances (35) in the HACT study cohorts, during acute COVID-19 OD co-occurred frequently with taste disorders, multiple non-specific infection symptoms, diminished appetite, rhinitis and sore throat (**Supplementary Figure S7**). During recovery, this association disappeared owing to the faster resolution of most of these acute infection symptoms. Importantly, persistent OD and hypogeusia were separated from other leading long COVID and PASC manifestations including tachypnea, fatigue and neurocognitive deficits (**Figure 3**).

**Figure 3.**
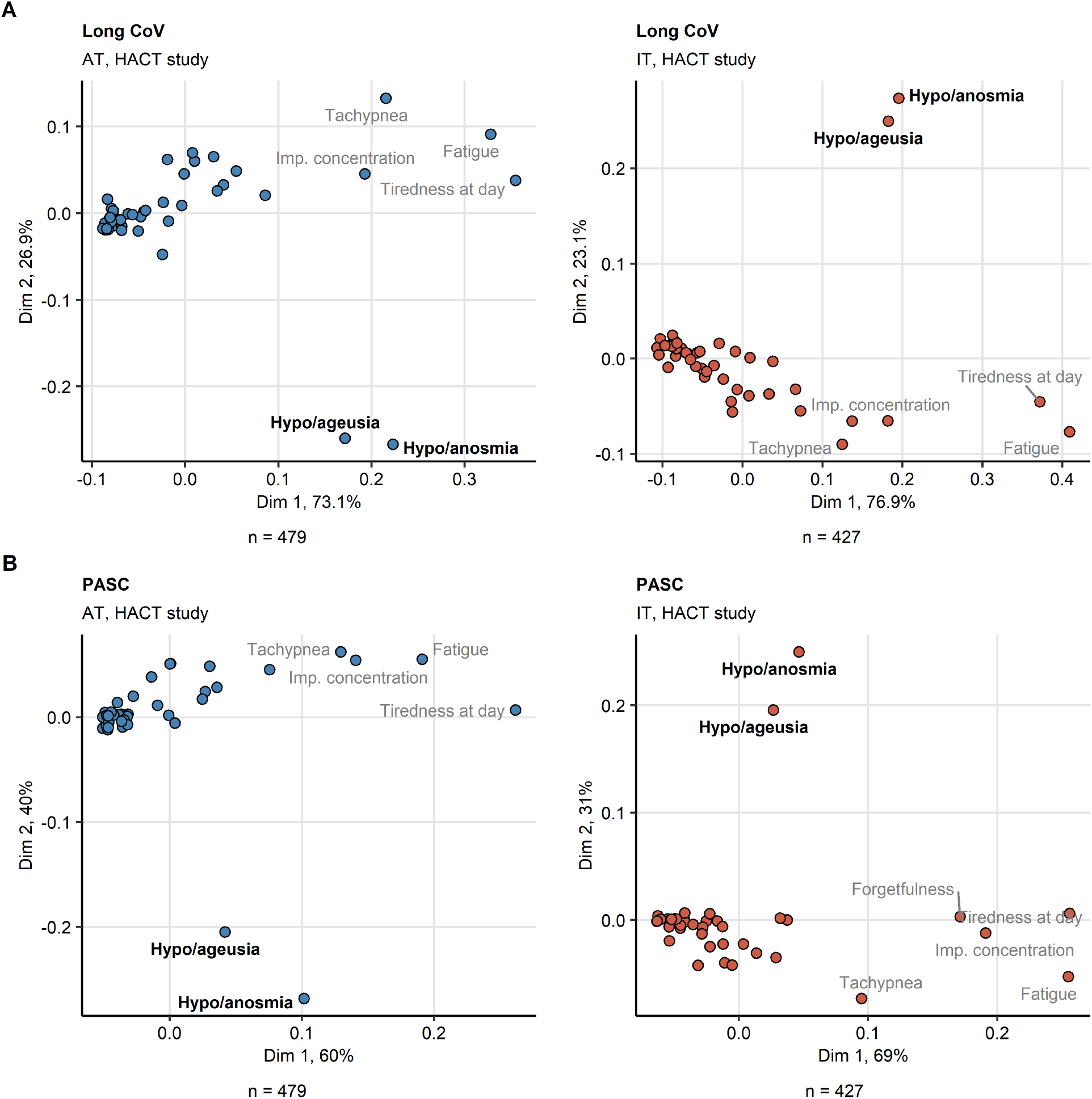
Self-reported smell and taste disorders are isolated symptoms of long COVID and PASC. Binary symptom occurrence data during for long COVID (≥ 28 days after clinical onset, **A**) and PASC (≥ 90 days, **B**) in the HACT Austria (AT) and Italy (IT) cohorts were subjected to two-dimensional multi-dimensional scaling (MDS) with simple matching distance (SMD) between the symptoms. MDS coordinates are presented in point plots. Selected data points are labeled with the symptom names. Percentages of the data set variance associated with the MDS dimensions are indicated in the plot axes. Numbers of complete observations are indicated under the plots. Imp.: impaired.

As investigated by association rule mining (37), roughly 90% of convalescents affected by taste disorders during the post-acute recovery suffered also from subjective OD (confidence for taste disorder → OD, long COVID: >91%, PASC: >88%). Furthermore, the concomitant self-reported hyposmia and hypogeusia were identified as highly frequent symptom pair in long COVID and PASC (support statistic, long COVID: >30%, PASC: >20% participants) (**Figure 4**, S**upplementary Tables S3** – **S4**).

**Figure 4.**
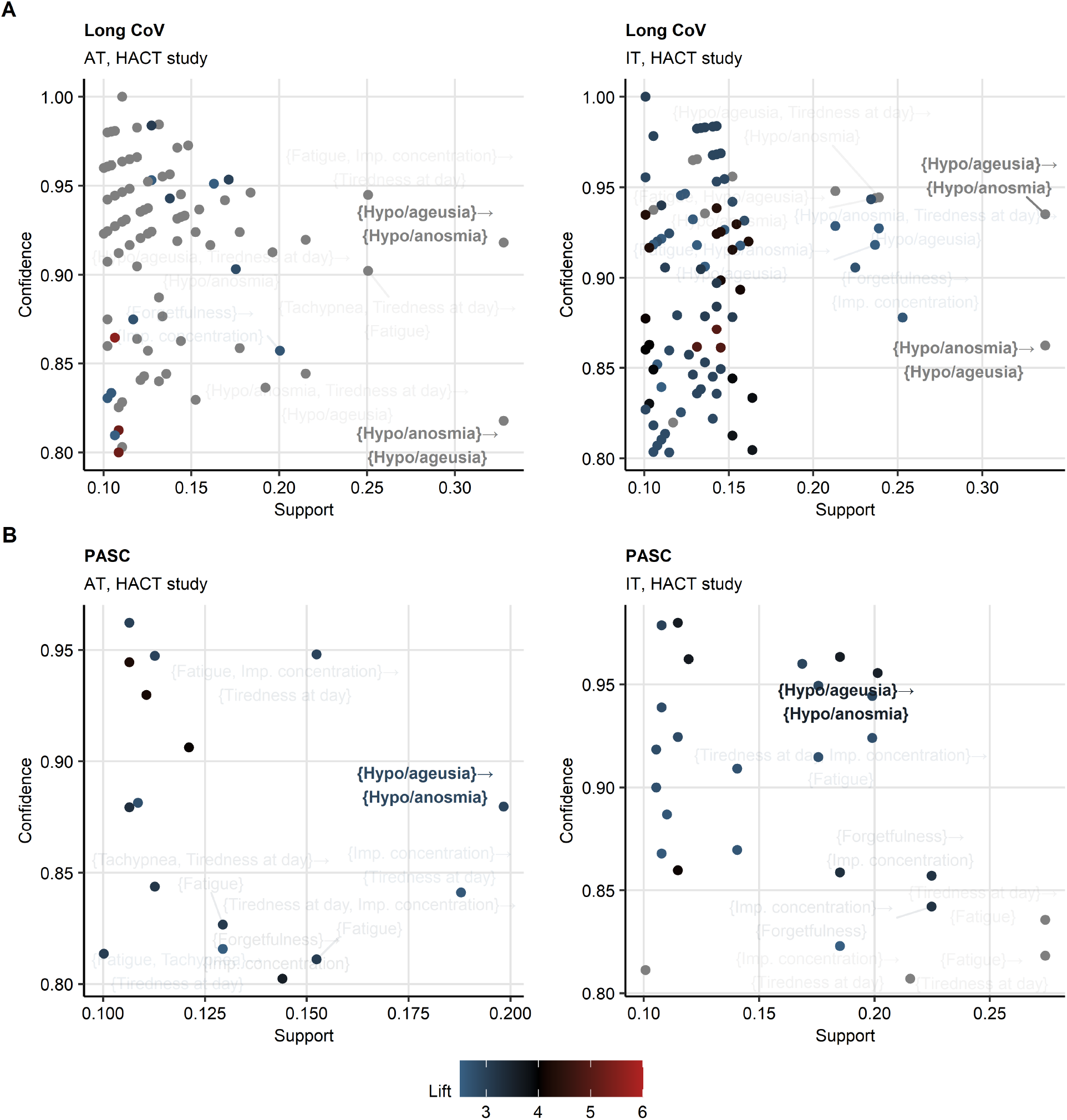
Frequent co-occurrence of smell and taste disorders in long COVID and PASC. Frequent combinations of symptoms of long COVID (**A**) and PASC (**B**) in the HACT study Austria (AT) and Italy (IT) cohorts were identified with the apriori algorithm. Symptom combination support and confidence are presented in the plots. Point color codes for lift values. The hypo/anosmia and hypo/ageusia combinations are labeled.

### Smell and taste disorder phenotype of COVID-19 recovery in the HACT study

By PAM (partitioning around medoids) semi-supervised clustering (38,40) of the HACT cohorts by the symptom-specific resolution times, three convalescent subsets were identified, termed further ‘COVID-19 recovery clusters’ (**Figure 5, Supplementary Figure S8AB**). Of note, tiredness, shortness of breath, self-reported concentration and memory impairments and self-reported OD were the most important clustering factors identified by permutation analysis (**Supplementary Figure S8C**).

**Figure 5.**
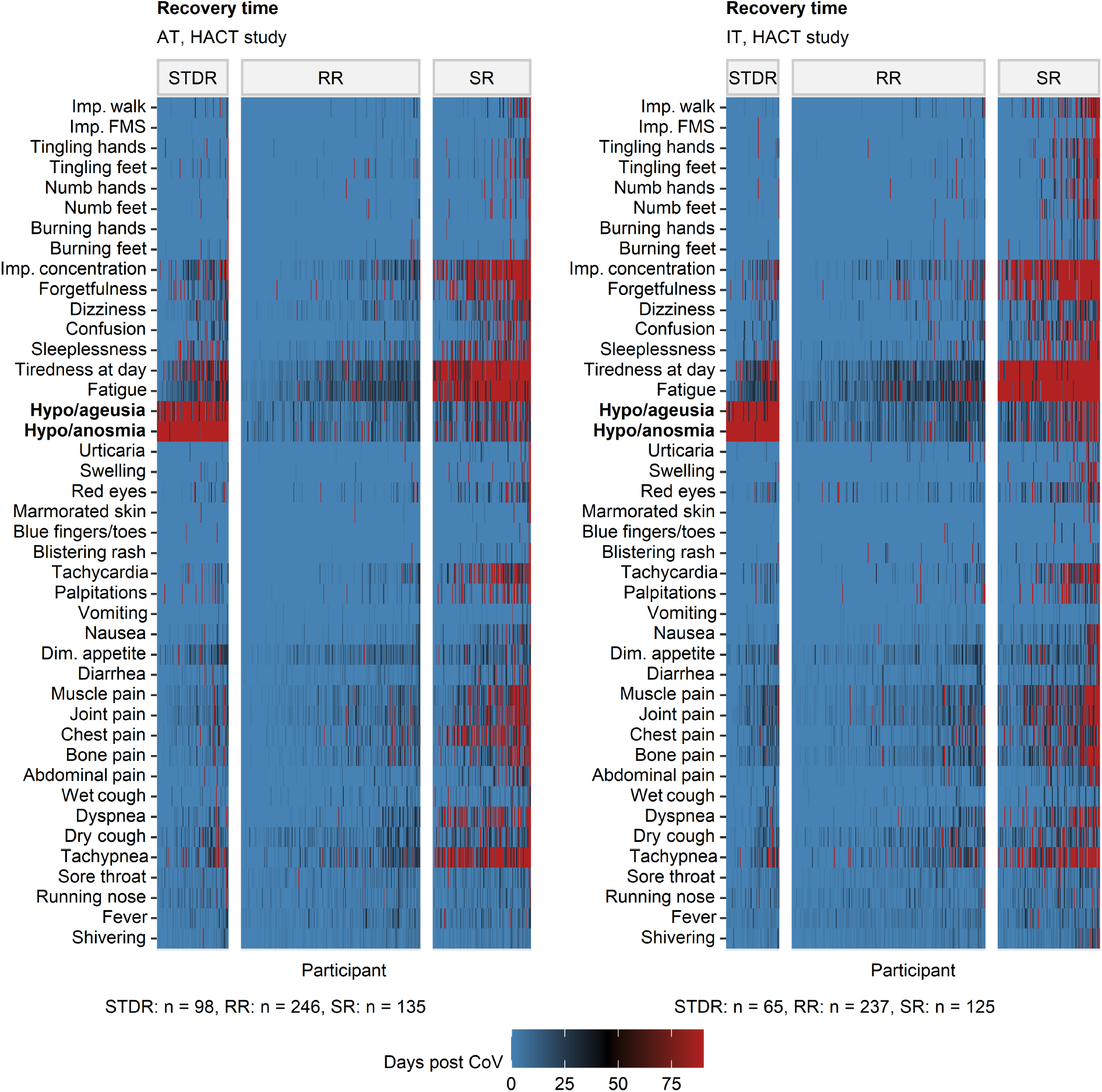
Clustering of ambulatory COVID-19 individuals by symptom-specific recovery times. Individuals of the training Austria (AT) cohort of the HACT study were subjected to clustering in respect to symptom-specific recovery times with the PAM (partitioning around medoids) algorithm and Euclidean distance measure (**Supplementary Figure S8**). Cluster assignment in the test Italy (IT) HACT cohort was done with k-NN label propagation algorithm. Recovery times for particular COVID-19 symptoms in the COVID-19 recovery clusters are presented as heat maps. Numbers of individuals assigned to the recovery clusters are indicated under the plots. Dim.: diminished, Imp.: impaired, FMS: fine motor skills.

The smell and taste disorder recovery cluster (STDR, AT: 21%, IT: 15% participants) was characterized by self-reported OD and taste disorders at day 90 of convalescence in almost all participants. Most other symptoms including tiredness, fatigue, respiratory and neurocognitive complaints resolved rapidly in the STDR cluster. The largest rapid-recovery (RR) cluster included >50% of the participants (AT: 51%, IT: 56%) and was characterized by both an oligosymptomatic acute COVID-19 course and rapid symptom resolution. By contrast, the slow-recovery (SR) cluster comprising up to 30% of the convalescents (AT: 28.2%, IT: 29.3%) was characterized by the highest number of acute and persistent complaints and particularly slow resolution of fatigue, tiredness, tachypnea and self-reported memory and concentration impairment (**Figure 5, Figure 6, Supplementary Figure S9 – S12)**.

**Figure 6.**
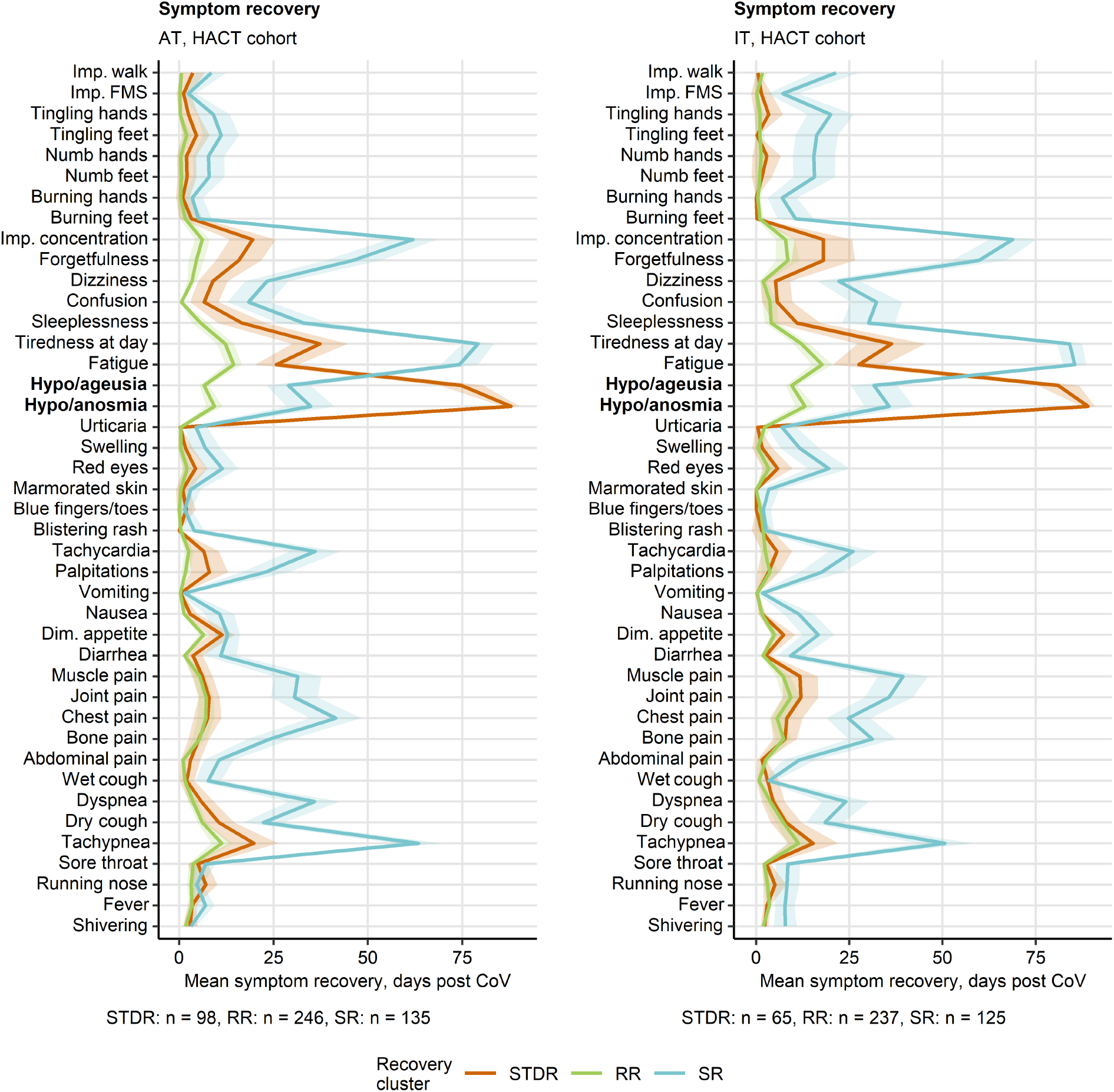
Duration of neurocognitive and respiratory symptoms, fatigue, smell and taste disorders differs between the COVID-19 recovery clusters. Semi-supervised clustering was performed as presented in **Figure 5** and **Supplementary Figure S8**. Mean recovery times in the recovery clusters are presented as lines, 2 *×* SEM intervals are displayed as tinted regions. Numbers of individuals assigned to the recovery clusters are indicated under the plots. Dim.: diminished, Imp.: impaired, FMS: fine motor skills.

Concerning demographic and clinical background, the severely affected SR cluster included the oldest participants with the highest comorbidity and daily medication rates. The RR and STDR clusters had a similar comorbidity fraction but differed in the sex distribution with the highest percentage of females (AT: 79%, IT: 83%) in the STDR cluster (**Figure 7, Supplementary Tables S5** – **S6**).

**Figure 7.**
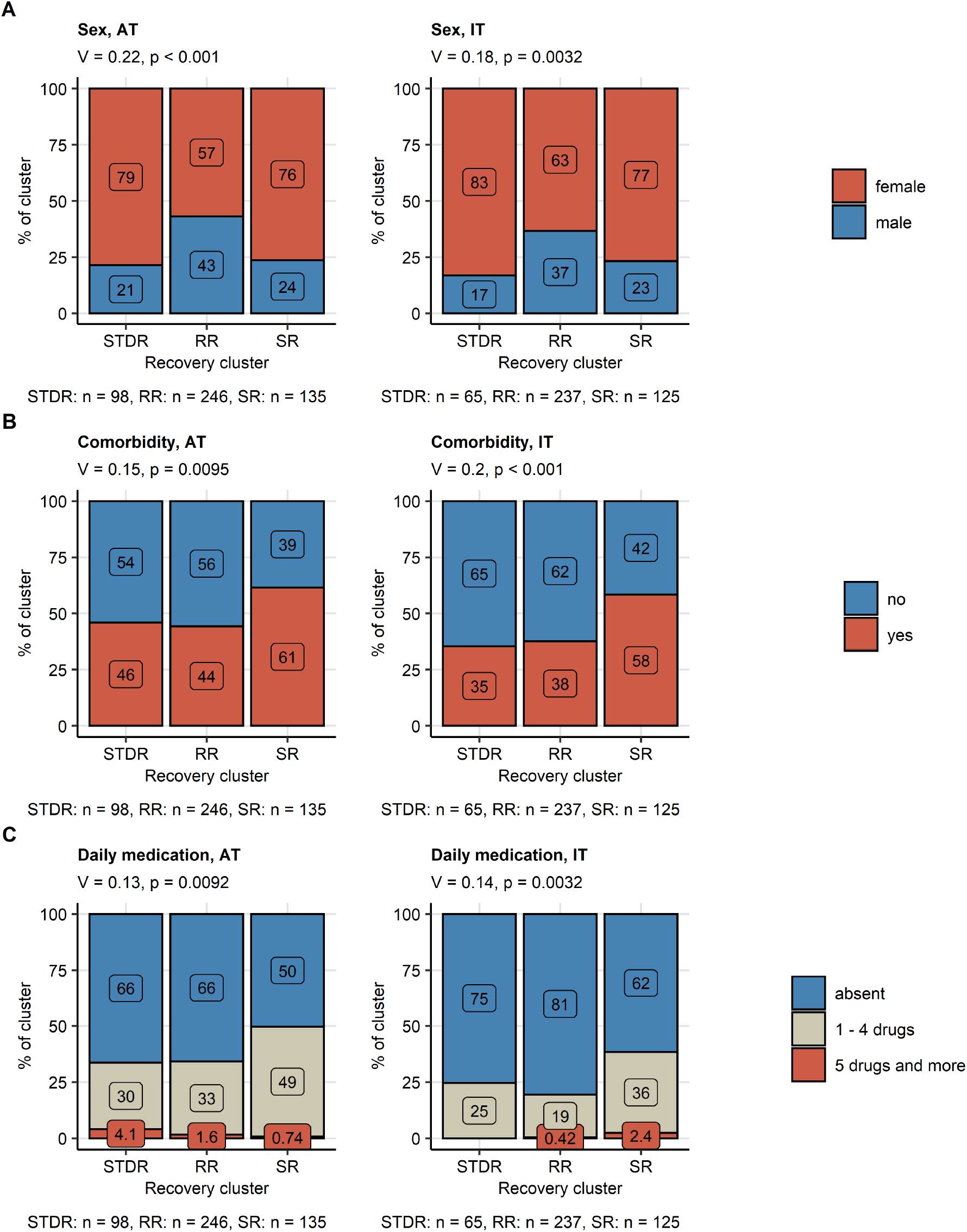
COVID-19 recovery clusters differ in sex distribution, comorbidity and daily medication rates. Differences in sex distribution (**A**), frequency of comorbidity (**B**) and daily medication (**C**) between the recovery clusters in the training Austria (AT) and the test Italy (IT) HACT study cohorts (**Figure 5, Supplementary Figure S8**) were assessed by *χ*^2^ test with Cramer V effect size statistic. P values were corrected for multiple testing with Benjamini-Hochberg method. The frequencies are presented as bar plots. Effect size statistics and p values are indicated in the plot caption. Numbers of complete observations are displayed under the plots.

The RR and STDR cluster individuals reported comparable frequencies of weight loss, new drugs, need of rehabilitation and loss of physical performance at the survey completion. In comparison, those rates were significantly elevated in the SR cluster. Furthermore, the RR and the STDR cluster individuals displayed the lowest scoring of anxiety, depression, mental stress, self-reported mental health impairment and quality of life loss. By contrast, the worst readouts for mental health and quality of life status at survey completion were observed in the SR cluster (**Figure 8, Supplementary Figure S13** – **S14, Supplementary Tables S7 – S8**).

**Figure 8.**
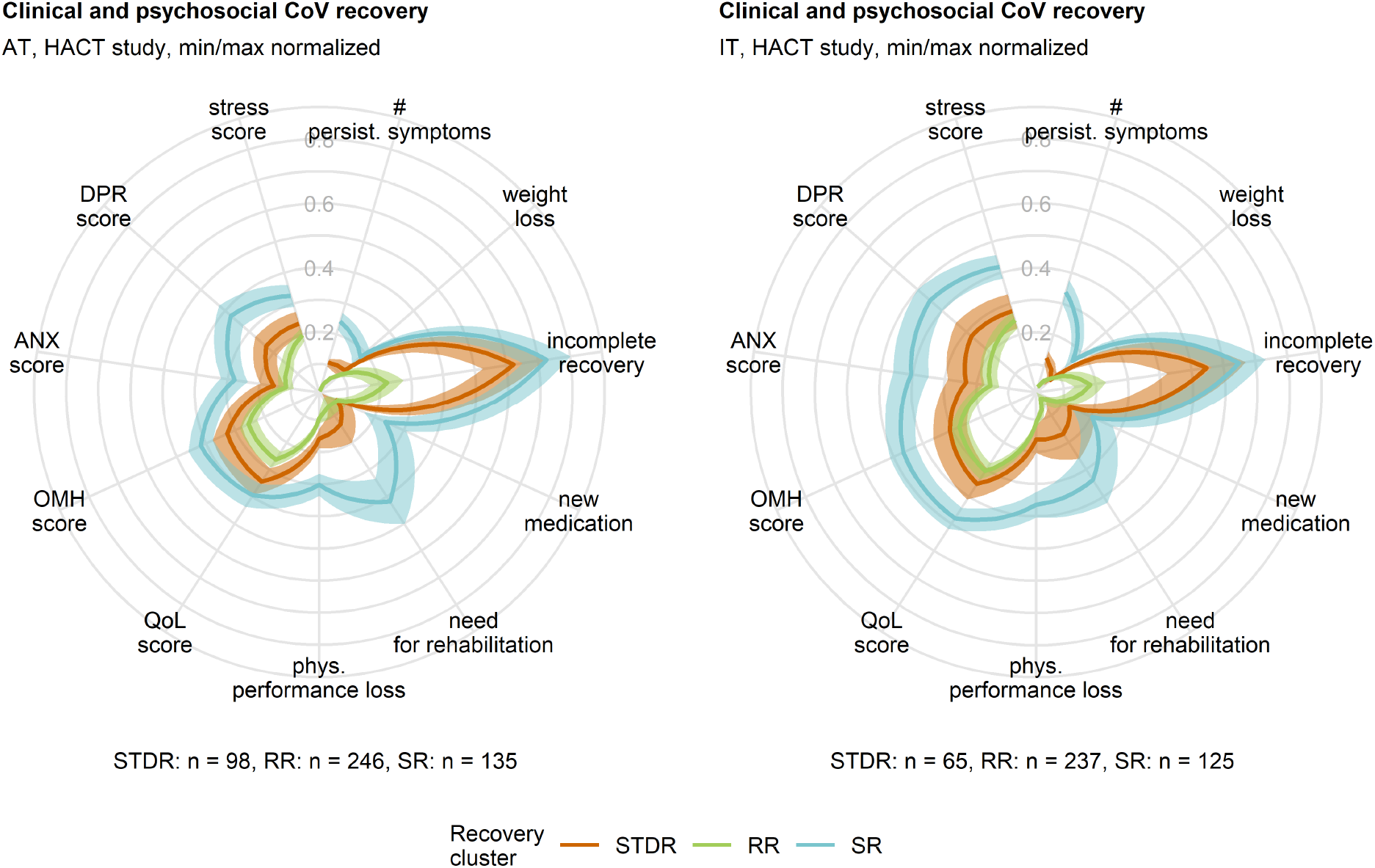
Good clinical, mental and psychosocial recovery at the survey completion in individuals with persistent smell and taste disorders. Differences in minimum/maximum scaled readouts of clinical and physical recovery and psychosocial health scoring in the recovery clusters (**Figure 5, Supplementary Figure S8**) are presented in radial plots. yes/no items (incomplete convalescence, weight loss, new medication and need for rehabilitation) were binarized (yes: 1, no: 0) prior to visualization. Lines represent mean values, 2 *×* SEM intervals are displayed as tinted regions. Numbers of individuals assigned to the recovery clusters are indicated under the plots. Incomplete recovery: self-reported incomplete recovery, # persist. symptoms: number of long COVID symptoms (≥ 28 days after clinical onset), phys. performance loss: physical performance loss as compared with the time before COVID-19, QoL score: quality of life impairment score, OMH score: overall mental health impairment score, ANX score: anxiety score, DPR: depression score.

## Discussion

We demonstrated that OD is frequent in hospitalized and non-hospitalized COVID-19 patients. Slow recovery of self-perceived OD was accompanied by taste disorders and independent of other common post-acute COVID-19 sequelae such as fatigue, tiredness, tachypnea or neurocognitive deficits. Convalescents with persistent OD were predominantly young females with low comorbidity rates and were characterized by good mental health and high ratings of physical performance at the time of study participation, i. e ≥90 days after COVID-19 onset. These findings may qualify persistent smell and taste disorders as a distinct phenotype of long COVID and PASC.

In our analysis, self-perceived OD was a common acute symptom of ambulatory (47 – 75% patients) and moderate-to-severe COVID-19 (33 – 53%), as described before (2,7,16,20) Percentages of self-perceived OD halved during the first four weeks of convalescence. Still, every sixth mild or moderate COVID-19 patient of the CovILD cohort reported persistent OD at the one-year follow-up. This recovery kinetics resembles the previously reported fast OD resolution within the first few weeks followed by a plateau (19,42). Since data beyond one year follow-up are lacking, it is still unclear whether post-COVID-19 OD resolves in all patients (43). As observed in other viral diseases or traumatic brain injury (TBI), complete resolution may take months to years which may leave millions of COVID-19 convalescents worldwide with residual OD during the pandemic (44–46). Hence, there is a huge need to develop novel therapeutic approaches. It was demonstrated that olfactory training may significantly ameliorate OD in viral diseases or TBI individuals (44) and remains the recommended first line treatment in post-COVID-19 OD (47–50).

The apparently higher rate of objective OD compared to self-perceived hyposmia in the CovILD cohort may be partially explained by the presence of objective COVID-19-independent OD, whose prevalence is estimated to be as high as 29% in the general population (51). In addition, the effect of age on olfactory function in the CovILD cohort cannot be excluded (51). Although our observations suggest that the rate of self-reported hyposmia underestimates the objective post-COVID-19 OD frequency, especially in severe disease and long-term recovery, more research is needed to clarify that.

Our results corroborate that recovery times of different COVID-19 symptoms vary significantly (1,7). Most respiratory and non-specific infection manifestations resolved within 14 days after COVID-19 onset. Yet, substantially slower recovery was observed for OD, taste disorders, tiredness, fatigue and neurocognitive symptoms being therefore the leading manifestations of long COVID and PASC.

Phenotypes or patterns of co-existing manifestations of long COVID and PASC are being gradually recognized by recent literature (1,6,7,9,52) and are crucial to predict individual outcomes and tailor specific rehabilitation needs. We found that post-acute subjective OD frequently co-occurred with self-reported taste disorders, which likely reflect an impaired retronasal smell (19), and had little overlap with other persistent complaints, especially in the period ≥90 days after COVID-19 onset. Our clustering results suggest that the persistent smell and taste disorder recovery phenotype may be regarded as a distinct form of long COVID and PASC affecting preferentially younger, female individuals without pre-existing chronic conditions. Previous studies described particularly high rates of acute OD in this population, which may be extrapolated to the later course (53,54). Consequently, female patients with OD during acute COVID-19 may best benefit from early neurological and/or ENT assessment and timely initiated therapy.

Multiple previous studies linked post-COVID-19 OD with impaired quality of life, anxiety and depression (20,22–24). In our study, persistent OD affected predominantly a subset of patients without other clinical, physical, and mental health complaints. This discrepancy may be partly explained by methodological differences as we did not use questionnaires specifically addressing OD related QoL measures. More importantly, our results underline the differing impact of long COVID/PASC phenotypes on daily life functioning. In contrast to the smell and taste disorder phenotype, persistent fatigue, neurocognitive and respiratory sequelae as observed in the ‘slow-recovery phenotype’ were paralleled by impaired physical and mental health as well as decreased quality of life.

Our study bears some limitations. Females and health care workers were over-represented in the HACT collective and we only included patients with a symptomatic COVID-19 course indicating a selection bias towards health-aware long COVID-19 individuals. Short follow-up times and retrospective surveying of the symptom duration as pre-defined classes may have limited the analysis precision and precluded investigation of possible symptom relapses (1,6). In the CovILD cohort a possible dropout of participants with subjective complete recovery was likely a source of selection bias. Finally, we were not able to investigate effects of immunization, improved medication and the most recent SARS-CoV-2 variants in our study cohorts which were recruited predominantly the wild type and alpha variant outbreaks. In particular, OD following an omicron-variant SARS-CoV-2 infection is reportedly less frequent as compared with the wild type, alpha or delta pathogen (12,13). Hence, there is a continuous need for phenotyping of COVID-19-related OD as new variants of concern emerge.

## Conclusion

Our multi-cohort analysis describes slow-pace resolution of subjective OD both in COVID-19 inpatients and outpatients. Except for taste disorders, persistent OD was largely independent from other post-acute sequelae such as fatigue, tachypnea or neurocognitive manifestations. In contrast to a prolonged COVID-19 recovery phenotype, the OD phenotype was characterized by the absence of physical or mental health deficits. This stresses the heterogeneity of post-acute COVID-19 sequelae, each requiring tailored management strategies.

## Supporting information

STROBE checklist

## Data Availability

The raw data files will be made available upon request.

https://github.com/PiotrTymoszuk/hyposmia_analsis_pipeline

## Acknowledgments

We acknowledge commitment of the study participants, medical staff and public health service workers during the SARS-CoV-2 pandemic.

## Author’s contribution

VR and PT analyzed the data, VR, PT, JLR and RH wrote the manuscript, VR, SS, BH, DA, AL, MK, PM, AB, KH, AP, TS, KK, MP, PK, LP, EF, AD recorded the study data, VR, PT, SS, TS, JLR and RH curated the data, VR, SS, DA, AP, KH, TS, B. Pfeifer, SK, AD, AH, CJW, BSU, EW, RB, AJS, RBW, HB, IT, B. Pfausler, GP, KS, GW, JLR and RH designed and supervised the studies and participated in the result evaluation.

## Data and code availability

The raw data files will be made available upon request. The entire analysis pipeline was published at https://github.com/PiotrTymoszuk/hyposmia_analsis_pipeline.

