## Supplementary Material for "Distinct smell and taste disorder phenotype of post-acute COVID-19 sequelae"

HACT and CovILD study teams

### Supplementary Methods

#### Study procedures and variables

The complete list of study variables and stratification scheme is provided in **Supplementary Table S1** for the HACT study and in **Supplementary Table S2** for the CovILD study.

##### COVID-19 symptoms

A total of 42 was surveyed in the HACT study cohorts (**Supplementary Figure S1** and **Supplementary Table S1**). The symptom duration was coded as follows: absent: 0 days, 1 - 3 days: 3 days, up to 1 week: 7 days, up to 2 weeks: 14 days, up to 4 weeks: 28 days, up to 3 months: 90 days, up to 6 months: 90 days, over 6 months: 90 days. Acute symptoms were defined as complaints present during the first 14 days after clinical onset of COVID-19. Long COVID symptoms were defined as complaints present for ≥ 28 days. PASC symptoms were defined as manifestations present for ≥ 90 days (1,2).

In the CovILD study, a total of 8 symptoms (reduced physical performance, hyposmia/anosmia, dyspnea, sleep problems, cough, fever, night sweating, gastrointestinal symptoms) were prospectively recorded with a standardized questionnaire at each of 60-, 100-, 180- and 360-day post COVID-19 follow-up (**Supplementary Figure S3** and **Supplementary Table S2**). Acute COVID-19 symptoms were assessed retrospectively (3,4).

##### Rating of physical recovery, mental health and quality of life in the HACT study

Self-perceived complete recovery, rehabilitation need and new medication since COVID-19 at the time of study participation were surveyed as single yes/no items. Percentage of physical performance loss as compared with the time before COVID-19 was rated with a 0 - 100% scale (5,6). Quality of life impairment (QoL) and overall mental health impairment (OMH) were rated with a 4 item Likert scale each (possible answers: "excellent," "good," "fair," "poor," scored: 0, 1, 2, and 3) (5,6). Anxiety (ANX) and depression (DPR) were assessed with the Patient Health Questionnaire (PHQ-4) (5–7). Psychosocial stress was scored with a modified 7 item PHQ stress module as described before (5,6,8).

##### Rating of hyposmia with sniffing stick test

Objective hyposmia at the 100-day and 360-day follow-up in the CovILD study participants was investigated with the 16-item sniffing stick test as described (9). Clinically relevant hyposmia was defined as ≤ 12 correct answers (9,10). In the analysis, participants with the complete answers concerning self-reported hyposmia and complete test results were included.

### Statistical analysis

#### Data transformation, descriptive statistic

Data transformation and statistical analysis was done with R version 4.0.5 with *tidyverse* data science environment (11). Analysis results were visualized with *ggplot2* (12), *cowplot* (13) as well as in-house developed *ExDA* (https://github.com/PiotrTymoszuk/ExDA) and *figur* (https://github.com/PiotrTymoszuk/figur) packages.

Descriptive statistics including median with interquartile ranges and frequency of complete answers for numeric and categorical variables were calculated with base R functions and *ExDA* package.

#### Statistical hypothesis testing

Since multiple study variables were non-normally distributed as assessed by Shapiro-Wilk test and visual assessment of their distribution (quantile-quantile plots), statistical significance for differences in outcome numeric variables were assessed with Mann-Whitney U test by Wilcoxon r effect size statistic (two groups) or Kruskal-Wallis test with $\eta^2$ effect size statistic. Differences in frequency distribution for categorical outcome variables were assessed by $\chi^2$ test with Cramer V effect size statistic. Interrater assessment of self-reported and sniffing test hyposmia was accomplished with Cohen's $\kappa$ statistic (14). $\kappa$ significance ($\kappa \neq 0$) was estimated with Wald Z test. P values were adjusted for multiple testing with Benjamini-Hochberg method (15) separately for each analysis task and cohort. Packages *rstatix* (16), *vcd* (17) and *Exda* (https://github.com/PiotrTymoszuk/ExDA) were used for statistical hypothesis testing.

#### Modeling of symptom recovery kinetic

To model recovery kinetics for binary symptom variables (0: absent, 1: present), second-order mixed-effect logistic (categorical features) modeling was applied (packages: *lme4*, *lmerTest* and development package *kinet* [https://github.com/PiotrTymoszuk/kinet]) (18–20). Each model followed the general formula:

$$Response \sim time + time^2 + (1 \vee individual)$$

where $(1 \vee individual)$ indicates the random effect of the individual and $time$ and $time^2$ indicate the first- and second-order time effect terms. The first-order term estimate was interpreted as a measure of the recovery speed and the second-order term estimate was used to assess the plateau/rebound effect. Significance of the accuracy gain of the full second-order model compared with the nested null model was determined by likelihood ratio test (LRT) versus the nested first-order and null models, respectively. Likelihood ratio $\lambda$ statistic (full versus null model) was used as an effect size measure. Individuals from the HACT or CovILD studies with the complete longitudinal symptom record were included in the kinetic modeling tasks. Results of the kinetic modeling were adjusted for multiple comparisons with Benjamini-Hochberg method (15).

#### Symptom-symptom distances and multi-dimensional scaling

To assess co-occurrence or exclusivity of symptoms, simple matching distances between manifestations of each acute COVID-19, long COVID and PASC in the HACT study cohorts were calculated (package *scrime* and development package *clustTools* [https://github.com/PiotrTymoszuk/clustTools]) (21,22). Subsequently, the distance matrix was subjected to multi-dimensional scaling (MDS, k = 2 dimensions, package *stats*, function *cmdscale()*). Association of specific symptoms was assessed by visual analysis of MDS coordinate plots.

##### Apriori analysis of acute COVID-19, long COVID and PASC symptoms in the HACT study

Frequent combinations of symptoms of long COVID and PASC in the HACT study cohorts were identified with the apriori algorithm (package *arules*) (23,24) with the minimal support cutoff of 0.1, 2 - 10 item transaction length, confidence > 0.8 and lift > 2. The support statistic were used to estimate the symptom combination frequency. The confidence value was treated as an estimate of conditional probability of the symptom co-occurrence. The lift statistic was interpreted as a measure of the symptom dependence (lift = 1, symptoms are independent). The complete analysis results are presented in **Supplementary Tables S3** and **S4**.

##### Clustering analysis

COVID-19 recovery clusters of the training Autria (AT) HACT cohort participants in respect to symptom-specific recovery times (**Figure 1A**) were defined with the PAM (partitioning around medoids) algorithm and Euclidean distance statistic (packages *cluster*, *philentropy* and development package *clustTools* [https://github.com/PiotrTymoszuk/clustTools]) (25,26). The set of participants with the complete clustering variable set was included in the analysis. The symptom recovery times were not subjected to any type of pre-processing. The choice of the clustering procedure was motivated by the analysis of the clustering variance (ratio of the total between-cluster to total sum of squares) and clustering structure stability in 10-fold cross-validation (metric: rate of correct cluster assignment, cluster assignment predicted by k = 5 nearest neighbors label propagation algorithm, package *clustTools*) (27,28) for several clustering algorithms as presented in **Supplementary Figure S8A**. The optimal number of clusters was determined by the bend of the total within-cluster sum of squares curve (**Supplementary Figure S8B**, package *factoextra*) (29). Permutation importance of specific clustering variables was investigated by calculating difference in clustering variance (ratio of total between-cluster sum of squares to total sum of squares) between the initial clustering object and the clustering object with the given variable reshuffled at random (package *clustTools*). Assignment of the

Italy HACT cohort participants to the recovery clusters was accomplished with k-nearest neighbors label propagation algorithm (k = 5) (28). The clustering efficacy in the training AT cohort and the test IT cohort measured by clustering variance statistic defined above was similar (AT: 0.59, IT: 0.57).

#### Data and source code availability

The raw data files will be made available upon request. The entire analysis pipeline was published at https://github.com/PiotrTymoszuk/hyposmia_analsis_pipeline.

#### Supplementary Tables

**Supplementary Table S1:** HACT study variables.

| Variable name | Variable label | Unit | Description |
|---|---|---|---|
| ID | Patient ID | | patient ID |
| cohort | Cohort | | Study cohort |
| acute_covid | Acute COVID-19 symptoms | | Acute COVID-19 symptoms (first two weeks) |
| sex | Sex | | participant's sex |
| age | Age | years | Participant's age at survey completion |
| bmi_class_before | BMI before COVID-19 | | Body mass index class before COVID-19 |
| cohabitants | Household size | persons | Household size in persons |
| household_size | Household size | | Household size class: single, pair or bigger |
| education_class | Education | | Tertiary vs. non-tertiary education |
| employment_before | Employment status | | Employment status before COVID-19 |
| employment_sector | Employment sector | | Employment sector |
| obs_time | Observation time | | Observation time: test to survey completion |
| smoking | Smoking history | | Smoking history before COVID-19 |
| comorb_present | Comorbidity | | At least one comorbidity present |
| comorb_sum | Sum of co-morbidities | | Sum of the co-morbidities queried in the survey |
| hypertension | Hypertension | | Hypertension before acute COVID-19 |
| heart_circulation | Cardiovascular disease | | Cardiovascular disease before acute COVID-19 |
| diabetes | Diabetes | | Diabetes before acute COVID-19 |
| lung | Pulmonary disease | | Pulmonary disease before COVID-19 |
| gastrointenstinal | Gastrointestinal disease | | Gastrointestinal disease before COVID-19 |
| malignancy | Malignancy | | Malignancy before acute COVID-19 |
| hay_fever | Hay fever/allergy | | Hay fever or allergy before acute COVID-19 |

| Variable name | Variable label | Unit | Description |
|---|---|---|---|
| autoimmunity | Autoimmunity | | Autoimmune disease before acute COVID-19 |
| frequent_flu_like | Freq. resp. infections | | Frequent upper respiratory tract infections (more than two per year) before COVID-19 |
| two_plus_infections_antibiotics | Freq. bact. Infections | | Frequent bacterial infections requiring antibiotic treatment (more than two per year) before COVID-19 |
| depression_burnout | Pre-CoV depression/anxiety | | Depression, anxiety or burnout before acute COVID-19 |
| insomnia | Pre-CoV sleep disorders | | Sleep disorders before COVID-19 |
| night_dyspnoe | Sleep apnea | | Night apnea before COVID-19 |
| bruxism | Bruxism | | Bruxism before COVID-19 |
| pins_needles_feet | Feet paresthesia | | Feet/leg paresthesia before acute COVID-19 |
| daily_medication | Daily medication | | Daily medication class, number of drugs taken daily before COVID-19 |
| cov_outbreak | SARS-CoV2 outbreak | | Infection during the spring 2020, summer/fall 2020, winter/spring 2021 |
| illness_feeling | Severe illness feeling | | Subjective feeling of acute infection |
| sum_symptoms_acute | # acute symptoms | | Sum of acute symptoms (first two weeks) |
| sum_symptoms_long | # persistent symptoms | | Sum of persistent symptoms (4 weeks and longer) |
| weight_loss_kg | Weight loss | kg | Weight loss during/after acute COVID-19 |
| hair_loss | Hair loss | | Hair loss during/after acute COVID-19 |
| incomplete_covelescence | Incomplete recovery | | Self-perceived incomplete convalescence |
| perf_impairment | Physical performance loss | percent | Percent loss of physical performance after COVID-19 as compared with the time before |
| new_medication_fup | New medication after COVID-19 | | New medication after COVID-19 |
| rehabilitation_fup_needed | Subjective need for rehabilitation | | Self-reported need for rehabilitation after COVID-19 |
| phq_anxiety_score | ANX score | | Anxiety score |

| Variable name | Variable label | Unit | Description |
|---|---|---|---|
| phq_depression_score | DPR score | | Depression score |
| stress_score | Stress score | | Stress score |
| mental_health_score | OMH score | | Overall mental health score |
| life_quality_score | QoL score | | Quality of life score |
| fever | Fever | | Fever |
| ague | Shivering | | Shivering |
| sore_throat | Sore throat | | Sore throat |
| running_nose | Running nose | | Running nose |
| fatigue | Fatigue | | Fatigue |
| dry_cough | Dry cough | | Dry cough |
| wet_cough | Wet cough | | Wet cough |
| breath_short | Tachypnea | | Tachypnea |
| dyspnoe | Dyspnea | | Dyspnea |
| chest_pain | Chest pain | | Chest pain |
| tachycardia | Tachycardia | | Tachycardia |
| extrasystole | Palpitations | | Palpitations |
| joint_pain | Joint pain | | Joint pain |
| bone_pain | Bone pain | | Bone pain |
| muscle_pain | Muscle pain | | Muscle pain |
| abdominal_pain | Abdominal pain | | Abd. pain |
| nausea | Nausea | | Nausea |
| vomiting | Vomiting | | Vomiting |
| dim_appetite | Dim. appetite | | Dim. appetite |
| diarrhea | Diarrhea | | Diarrhea |
| dizziness | Dizziness | | Dizziness |
| anosmia | Hypo/anosmia | | Hyposmia/anosmia |

| Variable name | Variable label | Unit | Description |
|---|---|---|---|
| taste_loss | Hypo/ageusia | | Hypogeusia/ageusia |
| confusion | Confusion | | Confusion |
| tingle_feet | Tingling feet | | Tingling feet |
| tingle_hands | Tingling hands | | Tingling hands |
| ache_feet | Burning feet | | Burning feet |
| ache_hands | Burning hands | | Burning hands |
| numb_feet | Numb feet | | Numb feet |
| numb_hands | Numb hands | | Numb hands |
| unhandiness_walk | Imp. walk | | Imp. walk |
| unhandiness_micromotor | Imp. FMS | | Imp. f. m. s. |
| sleep_prob | Sleeplessness | | Sleeplessness |
| fatigue_day | Tiredness at day | | Tiredness at day |
| imp_concentration | Imp. concentration | | Imp. concentration |
| forgetfulness | Forgetfulness | | Forgetfulness |
| swelling | Swelling | | Swelling |
| blue_fingers | Blue fingers/toes | | Blue fingers/toes |
| urticaria | Urticaria | | Urticaria |
| blister_rash | Blistering rash | | Blistering rash |
| net_rash | Marmorated skin | | Bl. marm. skin. |
| red_eyes | Red eyes | | Red eyes |

**Supplementary Table S2:** CovILD study variables.

| Variable name | Variable label | Unit |
|---|---|---|
| ID | | |
| time_numeric | Time post diagnosis | days |
| sex | Sex | |
| age | Age | years |
| weight_class | Weight class | |
| comorb_present | Comorbidity present | % |
| no_comorb | # comorbidities | |
| endometabolic_comorb | Metabolic disease | % |
| hypertension_comorb | Hypertension | % |
| cardiovascular_comorb | CVD | % |
| diabetes_comorb | Diabetes | % |
| pulmonary_comorb | Pulmonary disease | % |
| gastro_comorb | GID | % |
| malingancy_comorb | Malignancy | % |
| immdef_comorb | Immune deficiency | % |
| cat_WHO | COVID-19 severity | |
| sleep_sympt | Sleep problems | % |
| dyspnoe_sympt | Dyspnea | % |
| cough_sympt | Cough | % |
| fever_sympt | Fever | % |
| night_sweat_sympt | Night sweat | % |
| gastro_sympt | Gastrointestinal | % |
| anosmia_sympt | Hypo/anosmia | % |
| fatigue_sympt | Reduced performance | % |

**Supplementary Table S3:** Results of apriori analysis for long COVID-19 symptoms in the HACT study . Top 10 most frequent symptom combinations (highest support value) in the AT (Austria) and IT (Italy) cohort are presented. The full results are available as a supplementary Excel table.

| Cohort | Symptoms | Support | Confidence | Lift |
|---|---|---|---|---|
| AT | {Tachypnea, Tiredness at day,Imp. concentration} → {Fatigue} | 0.184 | 0.946 | 2.10 |
| AT | {Fatigue, Hypo/anosmia} → {Hypo/ageusia} | 0.192 | 0.836 | 2.34 |
| AT | {Tachypnea, Imp. concentration} → {Fatigue} | 0.196 | 0.913 | 2.02 |
| AT | {Forgetfulness} → {Imp. concentration} | 0.200 | 0.857 | 2.67 |
| AT | {Hypo/ageusia, Tiredness at day} → {Hypo/anosmia} | 0.215 | 0.920 | 2.29 |
| AT | {Hypo/anosmia, Tiredness at day} → {Hypo/ageusia} | 0.215 | 0.844 | 2.36 |
| AT | {Fatigue, Imp. concentration} → {Tiredness at day} | 0.251 | 0.945 | 2.05 |
| AT | {Tachypnea, Tiredness at day} → {Fatigue} | 0.251 | 0.902 | 2.00 |
| AT | {Hypo/ageusia} → {Hypo/anosmia} | 0.328 | 0.918 | 2.29 |
| AT | {Hypo/anosmia} → {Hypo/ageusia} | 0.328 | 0.818 | 2.29 |
| IT | {Fatigue, Hypo/ageusia,Tiredness at day} → {Hypo/anosmia} | 0.213 | 0.948 | 2.42 |
| IT | {Fatigue, Hypo/anosmia,Tiredness at day} → {Hypo/ageusia} | 0.213 | 0.929 | 2.57 |
| IT | {Fatigue, Forgetfulness} → {Imp. concentration} | 0.225 | 0.906 | 2.72 |
| IT | {Tiredness at day, Forgetfulness} → {Imp. concentration} | 0.234 | 0.943 | 2.84 |
| IT | {Fatigue, Hypo/ageusia} → {Hypo/anosmia} | 0.237 | 0.944 | 2.41 |
| IT | {Fatigue, Hypo/anosmia} → {Hypo/ageusia} | 0.237 | 0.918 | 2.55 |
| IT | {Hypo/ageusia, Tiredness at day} → {Hypo/anosmia} | 0.239 | 0.944 | 2.41 |
| IT | {Hypo/anosmia, Tiredness at day} → {Hypo/ageusia} | 0.239 | 0.927 | 2.57 |
| IT | {Forgetfulness} → {Imp. concentration} | 0.253 | 0.878 | 2.64 |
| IT | {Hypo/ageusia} → {Hypo/anosmia} | 0.337 | 0.935 | 2.39 |
| IT | {Hypo/anosmia} → {Hypo/ageusia} | 0.337 | 0.862 | 2.39 |

**Supplementary Table S4:** Results of apriori analysis for PASC symptoms in the HACT study. Top 10 most frequent symptom combinations (highest support value) in the AT (Austria) and IT (Italy) cohort are presented. The full results are available as a supplementary Excel table.

| Cohort | Symptoms | Support | Confidence | Lift |
|---|---|---|---|---|
| AT | {Fatigue, Forgetfulness} → {Tiredness at day} | 0.113 | 0.947 | 2.95 |
| AT | {Tiredness at day, Forgetfulness} → {Fatigue} | 0.113 | 0.844 | 3.16 |
| AT | {Tiredness at day, Forgetfulness} → {Imp. concentration} | 0.121 | 0.906 | 4.06 |
| AT | {Fatigue, Tachypnea} → {Tiredness at day} | 0.129 | 0.816 | 2.54 |
| AT | {Tachypnea, Tiredness at day} → {Fatigue} | 0.129 | 0.827 | 3.09 |
| AT | {Forgetfulness} → {Imp. concentration} | 0.144 | 0.802 | 3.59 |
| AT | {Fatigue, Imp. concentration} → {Tiredness at day} | 0.152 | 0.948 | 2.95 |
| AT | {Tiredness at day, Imp. concentration} → {Fatigue} | 0.152 | 0.811 | 3.04 |
| AT | {Imp. concentration} → {Tiredness at day} | 0.188 | 0.841 | 2.62 |
| AT | {Hypo/ageusia} → {Hypo/anosmia} | 0.198 | 0.880 | 2.93 |
| IT | {Imp. concentration, Forgetfulness} → {Tiredness at day} | 0.185 | 0.823 | 2.51 |
| IT | {Tiredness at day, Forgetfulness} → {Imp. concentration} | 0.185 | 0.963 | 3.61 |
| IT | {Tiredness at day, Imp. concentration} → {Forgetfulness} | 0.185 | 0.859 | 3.27 |
| IT | {Tiredness at day, Imp. concentration} → {Fatigue} | 0.199 | 0.924 | 2.76 |
| IT | {Fatigue, Imp. concentration} → {Tiredness at day} | 0.199 | 0.944 | 2.88 |
| IT | {Hypo/ageusia} → {Hypo/anosmia} | 0.201 | 0.956 | 3.55 |
| IT | {Imp. concentration} → {Tiredness at day} | 0.215 | 0.807 | 2.46 |
| IT | {Forgetfulness} → {Imp. concentration} | 0.225 | 0.857 | 3.21 |
| IT | {Imp. concentration} → {Forgetfulness} | 0.225 | 0.842 | 3.21 |
| IT | {Tiredness at day} → {Fatigue} | 0.274 | 0.836 | 2.50 |
| IT | {Fatigue} → {Tiredness at day} | 0.274 | 0.818 | 2.50 |

**Supplementary Table S5:** Demographic and baseline clinical characteristic at the COVID-19 onset of the HACT study participants assigned to the recovery clusters, Austria (AT) cohort.

| Variable | Cluster #1 | Cluster #2 | Cluster #3 | Significance[a] | Effect size[a] |
|---|---|---|---|---|---|
| Sex | female: 79% (n = 77)<br>male: 21% (n = 21)<br>complete: n = 98 | female: 57% (n = 140)<br>male: 43% (n = 106)<br>complete: n = 246 | female: 76% (n = 103)<br>male: 24% (n = 32)<br>complete: n = 135 | p < 0.001 | V = 0.22 |
| Age, years | median: 42 [IQR: 30 - 50]<br>range: 21 - 80<br>complete: n = 98 | median: 43 [IQR: 29 - 53]<br>range: 18 - 77<br>complete: n = 246 | median: 48 [IQR: 38 - 53]<br>range: 21 - 70<br>complete: n = 135 | p = 0.045 | $\eta^2 = 0.012$ |
| BMI before COVID-19[b] | normal: 62% (n = 60)<br>overweight: 24% (n = 23)<br>obesity: 14% (n = 14)<br>complete: n = 97 | normal: 55% (n = 133)<br>overweight: 29% (n = 70)<br>obesity: 17% (n = 41)<br>complete: n = 244 | normal: 47% (n = 64)<br>overweight: 31% (n = 42)<br>obesity: 21% (n = 29)<br>complete: n = 135 | ns (p = 0.39) | V = 0.073 |
| Education | non-tertiary: 64% (n = 62)<br>tertiary: 36% (n = 35)<br>complete: n = 97 | non-tertiary: 63% (n = 154)<br>tertiary: 37% (n = 92)<br>complete: n = 246 | non-tertiary: 64% (n = 86)<br>tertiary: 36% (n = 49)<br>complete: n = 135 | ns (p = 0.99) | V = 0.012 |
| Employment status | employed: 87% (n = 85)<br>unemployed: 7.1% (n = 7)<br>leave: 3.1% (n = 3)<br>retired: 3.1% (n = 3)<br>complete: n = 98 | employed: 80% (n = 198)<br>unemployed: 9.3% (n = 23)<br>leave: 1.6% (n = 4)<br>retired: 8.5% (n = 21)<br>complete: n = 246 | employed: 85% (n = 115)<br>unemployed: 7.4% (n = 10)<br>leave: 0.74% (n = 1)<br>retired: 6.7% (n = 9)<br>complete: n = 135 | ns (p = 0.56) | V = 0.079 |
| Observation time | median: 180 [IQR: 130 - 210]<br>range: 93 - 400<br>complete: n = 98 | median: 190 [IQR: 130 - 220]<br>range: 90 - 400<br>complete: n = 246 | median: 180 [IQR: 140 - 220]<br>range: 90 - 380<br>complete: n = 135 | ns (p = 0.85) | $\eta^2 = -0.0029$ |
| Comorbidity | 46% (n = 45)<br>complete: n = 98 | 44% (n = 109)<br>complete: n = 246 | 61% (n = 83)<br>complete: n = 135 | p = 0.0095 | V = 0.15 |
| Hypertension | 9.2% (n = 9)<br>complete: n = 98 | 10% (n = 25)<br>complete: n = 246 | 13% (n = 17)<br>complete: n = 135 | ns (p = 0.82) | V = 0.041 |
| Cardiovascular disease | 0% (n = 0)<br>complete: n = 98 | 2.8% (n = 7)<br>complete: n = 246 | 2.2% (n = 3)<br>complete: n = 135 | ns (p = 0.36) | V = 0.076 |

| Variable | Cluster #1 | Cluster #2 | Cluster #3 | Significance[a] | Effect size[a] |
|---|---|---|---|---|---|
| Diabetes | 2% (n = 2)<br>complete: n = 98 | 1.6% (n = 4)<br>complete: n = 246 | 0.74% (n = 1)<br>complete: n = 135 | ns (p = 0.82) | V = 0.04 |
| Pulmonary disease | 0% (n = 0)<br>complete: n = 98 | 4.5% (n = 11)<br>complete: n = 246 | 5.2% (n = 7)<br>complete: n = 135 | ns (p = 0.14) | V = 0.1 |
| Gastrointestinal disease | 1% (n = 1)<br>complete: n = 98 | 2% (n = 5)<br>complete: n = 246 | 1.5% (n = 2)<br>complete: n = 135 | ns (p = 0.88) | V = 0.032 |
| Malignancy | 0% (n = 0)<br>complete: n = 98 | 0.81% (n = 2)<br>complete: n = 246 | 5.9% (n = 8)<br>complete: n = 135 | p = 0.0025 | V = 0.17 |
| Hay fever/allergy | 13% (n = 13)<br>complete: n = 98 | 17% (n = 41)<br>complete: n = 246 | 25% (n = 34)<br>complete: n = 135 | ns (p = 0.073) | V = 0.12 |
| Autoimmunity[c] | 7.1% (n = 7)<br>complete: n = 98 | 4.1% (n = 10)<br>complete: n = 246 | 11% (n = 15)<br>complete: n = 135 | ns (p = 0.056) | V = 0.12 |
| Freq. resp. infections[d] | 5.1% (n = 5)<br>complete: n = 98 | 5.3% (n = 13)<br>complete: n = 246 | 10% (n = 14)<br>complete: n = 135 | ns (p = 0.2) | V = 0.093 |
| Freq. bact. Infections | 1% (n = 1)<br>complete: n = 98 | 3.7% (n = 9)<br>complete: n = 246 | 9.6% (n = 13)<br>complete: n = 135 | p = 0.01 | V = 0.15 |
| Pre-CoV depression/anxiety | 6.1% (n = 6)<br>complete: n = 98 | 2.8% (n = 7)<br>complete: n = 246 | 9.6% (n = 13)<br>complete: n = 135 | p = 0.038 | V = 0.13 |
| Pre-CoV sleep disorders | 5.1% (n = 5)<br>complete: n = 98 | 2.4% (n = 6)<br>complete: n = 246 | 4.4% (n = 6)<br>complete: n = 135 | ns (p = 0.53) | V = 0.063 |
| Daily medication | absent: 66% (n = 65)<br>1 - 4 drugs: 30% (n = 29)<br>5 drugs and more: 4.1% (n = 4)<br>complete: n = 98 | absent: 66% (n = 162)<br>1 - 4 drugs: 33% (n = 80)<br>5 drugs and more: 1.6% (n = 4)<br>complete: n = 246 | absent: 50% (n = 68)<br>1 - 4 drugs: 49% (n = 66)<br>5 drugs and more: 0.74% (n = 1)<br>complete: n = 135 | p = 0.0092 | V = 0.13 |

[a]Categorical variables: $\chi^2$ test with Cramer V effect size statistic. Numeric variables: Kruskal-Wallis test with $\eta^2$ effect size statistic. P values corrected form multiple testing with Benjamini-Hochberg method.

[b]BMI: body mass index, overweight > 25 kg/m$^2$, obesity > 30 kg/m$^2$,

[c]Frequent respiratory infections, > 2 per year.

[d]Frequent bacterial infections with antibiotic therapy, > 2 per year.

**Supplementary Table S6:** Demographic and baseline clinical characteristic at the COVID-19 onset of the HACT study participants assigned to the recovery clusters, Italy (IT) cohort.

| Variable | Cluster #1 | Cluster #2 | Cluster #3 | Significance[a] | Effect size[a] |
|---|---|---|---|---|---|
| Sex | female: 83% (n = 54)<br>male: 17% (n = 11)<br>complete: n = 65 | female: 63% (n = 150)<br>male: 37% (n = 87)<br>complete: n = 237 | female: 77% (n = 96)<br>male: 23% (n = 29)<br>complete: n = 125 | p = 0.0032 | V = 0.18 |
| Age, years | median: 46 [IQR: 34 - 55]<br>range: 18 - 71<br>complete: n = 65 | median: 43 [IQR: 32 - 53]<br>range: 18 - 77<br>complete: n = 237 | median: 48 [IQR: 38 - 56]<br>range: 19 - 95<br>complete: n = 125 | p = 0.024 | $\eta^2 = 0.016$ |
| BMI before COVID-19[b] | normal: 82% (n = 53)<br>overweight: 12% (n = 8)<br>obesity: 6.2% (n = 4)<br>complete: n = 65 | normal: 67% (n = 155)<br>overweight: 26% (n = 61)<br>obesity: 6.5% (n = 15)<br>complete: n = 231 | normal: 57% (n = 70)<br>overweight: 28% (n = 35)<br>obesity: 15% (n = 18)<br>complete: n = 123 | p = 0.0077 | V = 0.14 |
| Education | non-tertiary: 57% (n = 37)<br>tertiary: 43% (n = 28)<br>complete: n = 65 | non-tertiary: 57% (n = 135)<br>tertiary: 43% (n = 102)<br>complete: n = 237 | non-tertiary: 62% (n = 78)<br>tertiary: 38% (n = 47)<br>complete: n = 125 | ns (p = 0.6) | V = 0.05 |
| Employment status | employed: 80% (n = 52)<br>unemployed: 11% (n = 7)<br>leave: 3.1% (n = 2)<br>retired: 6.2% (n = 4)<br>complete: n = 65 | employed: 79% (n = 188)<br>unemployed: 11% (n = 27)<br>leave: 2.5% (n = 6)<br>retired: 6.8% (n = 16)<br>complete: n = 237 | employed: 86% (n = 108)<br>unemployed: 4.8% (n = 6)<br>leave: 0% (n = 0)<br>retired: 8.8% (n = 11)<br>complete: n = 125 | ns (p = 0.27) | V = 0.1 |
| Observation time | median: 140 [IQR: 120 - 280]<br>range: 92 - 370<br>complete: n = 65 | median: 140 [IQR: 110 - 260]<br>range: 90 - 390<br>complete: n = 237 | median: 140 [IQR: 120 - 300]<br>range: 90 - 380<br>complete: n = 125 | ns (p = 0.21) | $\eta^2 = 0.0045$ |
| Comorbidity | 35% (n = 23)<br>complete: n = 65 | 38% (n = 89)<br>complete: n = 237 | 58% (n = 73)<br>complete: n = 125 | p < 0.001 | V = 0.2 |
| Hypertension | 7.7% (n = 5)<br>complete: n = 65 | 6.8% (n = 16)<br>complete: n = 237 | 12% (n = 15)<br>complete: n = 125 | ns (p = 0.28) | V = 0.083 |
| Cardiovascular disease | 0% (n = 0)<br>complete: n = 65 | 3.4% (n = 8)<br>complete: n = 237 | 4% (n = 5)<br>complete: n = 125 | ns (p = 0.34) | V = 0.077 |

| Variable | Cluster #1 | Cluster #2 | Cluster #3 | Significance[a] | Effect size[a] |
|---|---|---|---|---|---|
| Diabetes | 0% (n = 0)<br>complete: n = 65 | 0% (n = 0)<br>complete: n = 237 | 0.8% (n = 1)<br>complete: n = 125 | ns (p = 0.34) | V = 0.075 |
| Pulmonary disease | 3.1% (n = 2)<br>complete: n = 65 | 2.1% (n = 5)<br>complete: n = 237 | 4% (n = 5)<br>complete: n = 125 | ns (p = 0.6) | V = 0.051 |
| Gastrointestinal disease | 0% (n = 0)<br>complete: n = 65 | 0.42% (n = 1)<br>complete: n = 237 | 1.6% (n = 2)<br>complete: n = 125 | ns (p = 0.37) | V = 0.071 |
| Malignancy | 6.2% (n = 4)<br>complete: n = 65 | 2.5% (n = 6)<br>complete: n = 237 | 5.6% (n = 7)<br>complete: n = 125 | ns (p = 0.28) | V = 0.083 |
| Hay fever/allergy | 7.7% (n = 5)<br>complete: n = 65 | 11% (n = 27)<br>complete: n = 237 | 15% (n = 19)<br>complete: n = 125 | ns (p = 0.34) | V = 0.076 |
| Autoimmunity[c] | 6.2% (n = 4)<br>complete: n = 65 | 4.6% (n = 11)<br>complete: n = 237 | 9.6% (n = 12)<br>complete: n = 125 | ns (p = 0.25) | V = 0.089 |
| Freq. resp. infections[d] | 0% (n = 0)<br>complete: n = 65 | 1.3% (n = 3)<br>complete: n = 237 | 8.8% (n = 11)<br>complete: n = 125 | p < 0.001 | V = 0.2 |
| Freq. bact. Infections | 0% (n = 0)<br>complete: n = 65 | 0.42% (n = 1)<br>complete: n = 237 | 3.2% (n = 4)<br>complete: n = 125 | ns (p = 0.071) | V = 0.12 |
| Pre-CoV depression/anxiety | 4.6% (n = 3)<br>complete: n = 65 | 3% (n = 7)<br>complete: n = 237 | 9.6% (n = 12)<br>complete: n = 125 | p = 0.044 | V = 0.13 |
| Pre-CoV sleep disorders | 3.1% (n = 2)<br>complete: n = 65 | 1.7% (n = 4)<br>complete: n = 237 | 11% (n = 14)<br>complete: n = 125 | p < 0.001 | V = 0.2 |
| Daily medication | absent: 75% (n = 49)<br>1 - 4 drugs: 25% (n = 16)<br>5 drugs and more: 0% (n = 0)<br>complete: n = 65 | absent: 81% (n = 191)<br>1 - 4 drugs: 19% (n = 45)<br>5 drugs and more: 0.42% (n = 1)<br>complete: n = 237 | absent: 62% (n = 77)<br>1 - 4 drugs: 36% (n = 45)<br>5 drugs and more: 2.4% (n = 3)<br>complete: n = 125 | p = 0.0032 | V = 0.14 |

[a]Categorical variables: $\chi^2$ test with Cramer V effect size statistic. Numeric variables: Kruskal-Wallis test with $\eta^2$ effect size statistic. P values corrected form multiple testing with Benjamini-Hochberg method.

[b]BMI: body mass index, overweight > 25 kg/m$^2$, obesity > 30 kg/m$^2$,

[c]Frequent respiratory infections, > 2 per year.

[d]Frequent bacterial infections with antibiotic therapy, > 2 per year.

**Supplementary Table S7:** COVID-19 course and recovery in the HACT study participants assigned to the recovery clusters, Austria (AT) cohort.

| Variable | Cluster #1 | Cluster #2 | Cluster #3 | Significance[a] | Effect size[a] |
|---|---|---|---|---|---|
| SARS-CoV2 outbreak | spring 2020: 55% (n = 54)<br>summer/fall 2020: 43% (n = 42)<br>winter/spring 2021: 2% (n = 2)<br>complete: n = 98 | spring 2020: 63% (n = 156)<br>summer/fall 2020: 35% (n = 87)<br>winter/spring 2021: 1.2% (n = 3)<br>complete: n = 246 | spring 2020: 53% (n = 71)<br>summer/fall 2020: 47% (n = 64)<br>winter/spring 2021: 0% (n = 0)<br>complete: n = 135 | ns (p = 0.16) | V = 0.09 |
| Severe illness feeling | 1 week: 52% (n = 51)<br>2 weeks: 26% (n = 25)<br>3 weeks: 8.2% (n = 8)<br>4 weeks: 8.2% (n = 8)<br>over 4 weeks: 6.1% (n = 6)<br>complete: n = 98 | 1 week: 69% (n = 170)<br>2 weeks: 19% (n = 46)<br>3 weeks: 6.9% (n = 17)<br>4 weeks: 4.1% (n = 10)<br>over 4 weeks: 1.2% (n = 3)<br>complete: n = 246 | 1 week: 24% (n = 33)<br>2 weeks: 22% (n = 30)<br>3 weeks: 14% (n = 19)<br>4 weeks: 12% (n = 16)<br>over 4 weeks: 27% (n = 37)<br>complete: n = 135 | $p < 0.001$ | V = 0.34 |
| # acute symptoms[b] | median: 15 [IQR: 11 - 18]<br>range: 2 - 30<br>complete: n = 98 | median: 11 [IQR: 8 - 15]<br>range: 1 - 28<br>complete: n = 246 | median: 20 [IQR: 16 - 23]<br>range: 4 - 42<br>complete: n = 135 | $p < 0.001$ | $\eta^2 = 0.26$ |
| # persistent symptoms[c] | median: 3 [IQR: 2 - 5]<br>range: 1 - 12<br>complete: n = 98 | median: 0 [IQR: 0 - 1]<br>range: 0 - 6<br>complete: n = 246 | median: 7 [IQR: 4 - 12]<br>range: 1 - 34<br>complete: n = 135 | $p < 0.001$ | $\eta^2 = 0.72$ |
| Weight loss, kg | median: 0.5 [IQR: 0 - 3]<br>range: 0 - 8<br>complete: n = 98 | median: 0 [IQR: 0 - 2.1]<br>range: 0 - 11<br>complete: n = 244 | median: 2 [IQR: 0 - 4.5]<br>range: 0 - 15<br>complete: n = 134 | $p < 0.001$ | $\eta^2 = 0.039$ |
| Hair loss | 19% (n = 19)<br>complete: n = 98 | 9.3% (n = 23)<br>complete: n = 246 | 30% (n = 41)<br>complete: n = 135 | $p < 0.001$ | V = 0.24 |
| Incomplete recovery[d] | 62% (n = 61)<br>complete: n = 98 | 22% (n = 55)<br>complete: n = 245 | 73% (n = 98)<br>complete: n = 135 | $p < 0.001$ | V = 0.47 |
| Physical performance loss, percent | median: 10 [IQR: 4 - 25]<br>range: 0 - 69<br>complete: n = 98 | median: 3.5 [IQR: 0 - 14]<br>range: 0 - 100<br>complete: n = 244 | median: 25 [IQR: 15 - 42]<br>range: 0 - 92<br>complete: n = 134 | $p < 0.001$ | $\eta^2 = 0.26$ |

| Variable | Cluster #1 | Cluster #2 | Cluster #3 | Significance[a] | Effect size[a] |
|---|---|---|---|---|---|
| New medication after COVID-19 | 7.1% (n = 7)<br>complete: n = 98 | 7.3% (n = 18)<br>complete: n = 246 | 24% (n = 32)<br>complete: n = 135 | $p < 0.001$ | V = 0.23 |
| Subjective need for rehabilitation | 13% (n = 13)<br>complete: n = 98 | 6.5% (n = 16)<br>complete: n = 246 | 42% (n = 56)<br>complete: n = 134 | $p < 0.001$ | V = 0.4 |
| ANX score[e] | median: 0 [IQR: 0 - 2]<br>range: 0 - 5<br>complete: n = 98 | median: 0 [IQR: 0 - 1]<br>range: 0 - 6<br>complete: n = 246 | median: 1.5 [IQR: 0 - 2]<br>range: 0 - 6<br>complete: n = 134 | $p < 0.001$ | $\eta^2 = 0.11$ |
| DPR score[f] | median: 1 [IQR: 0 - 2]<br>range: 0 - 6<br>complete: n = 98 | median: 0 [IQR: 0 - 2]<br>range: 0 - 6<br>complete: n = 246 | median: 2 [IQR: 1 - 3]<br>range: 0 - 6<br>complete: n = 135 | $p < 0.001$ | $\eta^2 = 0.15$ |
| Stress score | median: 3.5 [IQR: 2 - 6]<br>range: 0 - 19<br>complete: n = 98 | median: 3 [IQR: 1 - 5]<br>range: 0 - 16<br>complete: n = 246 | median: 5 [IQR: 3 - 9]<br>range: 0 - 16<br>complete: n = 135 | $p < 0.001$ | $\eta^2 = 0.064$ |
| OMH score[g] | median: 1 [IQR: 0 - 1]<br>range: 0 - 3<br>complete: n = 98 | median: 1 [IQR: 0 - 1]<br>range: 0 - 3<br>complete: n = 246 | median: 1 [IQR: 1 - 2]<br>range: 0 - 3<br>complete: n = 135 | $p < 0.001$ | $\eta^2 = 0.072$ |
| QoL score[h] | median: 1 [IQR: 1 - 1]<br>range: 0 - 3<br>complete: n = 98 | median: 1 [IQR: 0 - 1]<br>range: 0 - 3<br>complete: n = 246 | median: 1 [IQR: 1 - 2]<br>range: 0 - 3<br>complete: n = 135 | $p < 0.001$ | $\eta^2 = 0.052$ |

[a]Categorical variables: $\chi^2$ test with Cramer V effect size statistic. Numeric variables: Kruskal-Wallis test with $\eta^2$ effect size statistic. P values corrected form multiple testing with Benjamini-Hochberg method.

[b]Number of acute COVID-19 symptoms.

[c]Number of long COVID symptoms.

[d]Self-reported incomplete recovery.

[e]ANX: anxiety.

[f]DPR: depression.

[g]OMH score: overall mental health impairment score

[h]QoL score: quality of life impairment score

**Supplementary Table S8:** COVID-19 course and recovery in the HACT study participants assigned to the recovery clusters, Italy (AT) cohort.

| Variable | Cluster #1 | Cluster #2 | Cluster #3 | Significance[a] | Effect size[a] |
|---|---|---|---|---|---|
| SARS-CoV2 outbreak | spring 2020: 34% (n = 22)<br>summer/fall 2020: 65% (n = 42)<br>winter/spring 2021: 1.5% (n = 1)<br>complete: n = 65 | spring 2020: 28% (n = 67)<br>summer/fall 2020: 71% (n = 169)<br>winter/spring 2021: 0.42% (n = 1)<br>complete: n = 237 | spring 2020: 32% (n = 40)<br>summer/fall 2020: 68% (n = 85)<br>winter/spring 2021: 0% (n = 0)<br>complete: n = 125 | ns ($p = 0.55$) | $V = 0.062$ |
| Severe illness feeling | 1 week: 43% (n = 28)<br>2 weeks: 25% (n = 16)<br>3 weeks: 12% (n = 8)<br>4 weeks: 11% (n = 7)<br>over 4 weeks: 9.2% (n = 6)<br>complete: n = 65 | 1 week: 67% (n = 157)<br>2 weeks: 15% (n = 36)<br>3 weeks: 8.1% (n = 19)<br>4 weeks: 6.8% (n = 16)<br>over 4 weeks: 3% (n = 7)<br>complete: n = 235 | 1 week: 20% (n = 25)<br>2 weeks: 18% (n = 23)<br>3 weeks: 16% (n = 20)<br>4 weeks: 13% (n = 16)<br>over 4 weeks: 33% (n = 41)<br>complete: n = 125 | $p < 0.001$ | $V = 0.35$ |
| # acute symptoms[b] | median: 14 [IQR: 9 - 19]<br>range: 4 - 31<br>complete: n = 65 | median: 12 [IQR: 8 - 15]<br>range: 2 - 30<br>complete: n = 237 | median: 21 [IQR: 16 - 26]<br>range: 8 - 38<br>complete: n = 125 | $p < 0.001$ | $\eta^2 = 0.3$ |
| # persistent symptoms[c] | median: 2 [IQR: 2 - 4]<br>range: 1 - 11<br>complete: n = 65 | median: 0 [IQR: 0 - 1]<br>range: 0 - 7<br>complete: n = 237 | median: 8 [IQR: 5 - 12]<br>range: 2 - 27<br>complete: n = 125 | $p < 0.001$ | $\eta^2 = 0.67$ |
| Weight loss, kg | median: 0 [IQR: 0 - 2]<br>range: 0 - 5<br>complete: n = 65 | median: 0 [IQR: 0 - 2]<br>range: 0 - 8<br>complete: n = 236 | median: 2 [IQR: 0 - 4]<br>range: 0 - 15<br>complete: n = 124 | $p < 0.001$ | $\eta^2 = 0.049$ |
| Hair loss | 25% (n = 16)<br>complete: n = 65 | 9.7% (n = 23)<br>complete: n = 237 | 31% (n = 39)<br>complete: n = 125 | $p < 0.001$ | $V = 0.25$ |
| Incomplete recovery[d] | 55% (n = 35)<br>complete: n = 64 | 18% (n = 43)<br>complete: n = 235 | 65% (n = 81)<br>complete: n = 125 | $p < 0.001$ | $V = 0.45$ |
| Physical performance loss, percent | median: 10 [IQR: 1 - 20]<br>range: 0 - 58<br>complete: n = 65 | median: 5 [IQR: 0 - 16]<br>range: 0 - 60<br>complete: n = 231 | median: 29 [IQR: 20 - 50]<br>range: 0 - 93<br>complete: n = 124 | $p < 0.001$ | $\eta^2 = 0.29$ |

| Variable | Cluster #1 | Cluster #2 | Cluster #3 | Significance[a] | Effect size[a] |
|---|---|---|---|---|---|
| New medication after COVID-19 | 13% (n = 8)<br>complete: n = 63 | 8.2% (n = 19)<br>complete: n = 231 | 20% (n = 25)<br>complete: n = 123 | $p = 0.0092$ | V = 0.16 |
| Subjective need for rehabilitation | 17% (n = 11)<br>complete: n = 65 | 3.8% (n = 9)<br>complete: n = 235 | 34% (n = 42)<br>complete: n = 124 | $p < 0.001$ | V = 0.37 |
| ANX score[e] | median: 1 [IQR: 0 - 2]<br>range: 0 - 6<br>complete: n = 65 | median: 0 [IQR: 0 - 2]<br>range: 0 - 6<br>complete: n = 237 | median: 2 [IQR: 1 - 4]<br>range: 0 - 6<br>complete: n = 125 | $p < 0.001$ | $\eta^2 = 0.14$ |
| DPR score[f] | median: 2 [IQR: 0 - 2]<br>range: 0 - 6<br>complete: n = 65 | median: 1 [IQR: 0 - 2]<br>range: 0 - 6<br>complete: n = 236 | median: 2 [IQR: 2 - 4]<br>range: 0 - 6<br>complete: n = 125 | $p < 0.001$ | $\eta^2 = 0.16$ |
| Stress score | median: 4 [IQR: 1 - 6]<br>range: 0 - 13<br>complete: n = 65 | median: 3 [IQR: 1 - 6]<br>range: 0 - 14<br>complete: n = 235 | median: 6 [IQR: 4 - 9]<br>range: 0 - 15<br>complete: n = 125 | $p < 0.001$ | $\eta^2 = 0.11$ |
| OMH score[g] | median: 1 [IQR: 0 - 1]<br>range: 0 - 2<br>complete: n = 65 | median: 1 [IQR: 0 - 1]<br>range: 0 - 3<br>complete: n = 237 | median: 1 [IQR: 1 - 2]<br>range: 0 - 3<br>complete: n = 125 | $p < 0.001$ | $\eta^2 = 0.1$ |
| QoL score[h] | median: 1 [IQR: 1 - 1]<br>range: 0 - 3<br>complete: n = 65 | median: 1 [IQR: 0 - 1]<br>range: 0 - 3<br>complete: n = 237 | median: 1 [IQR: 1 - 2]<br>range: 0 - 3<br>complete: n = 125 | $p < 0.001$ | $\eta^2 = 0.095$ |

[a]Categorical variables: $\chi^2$ test with Cramer V effect size statistic. Numeric variables: Kruskal-Wallis test with $\eta^2$ effect size statistic. P values corrected form multiple testing with Benjamini-Hochberg method.

[b]Number of acute COVID-19 symptoms.

[c]Number of long COVID symptoms.

[d]Self-reported incomplete recovery.

[e]ANX: anxiety.

[f]DPR: depression.

[g]OMH score: overall mental health impairment score

[h]QoL score: quality of life impairment score

#### Supplementary Figures

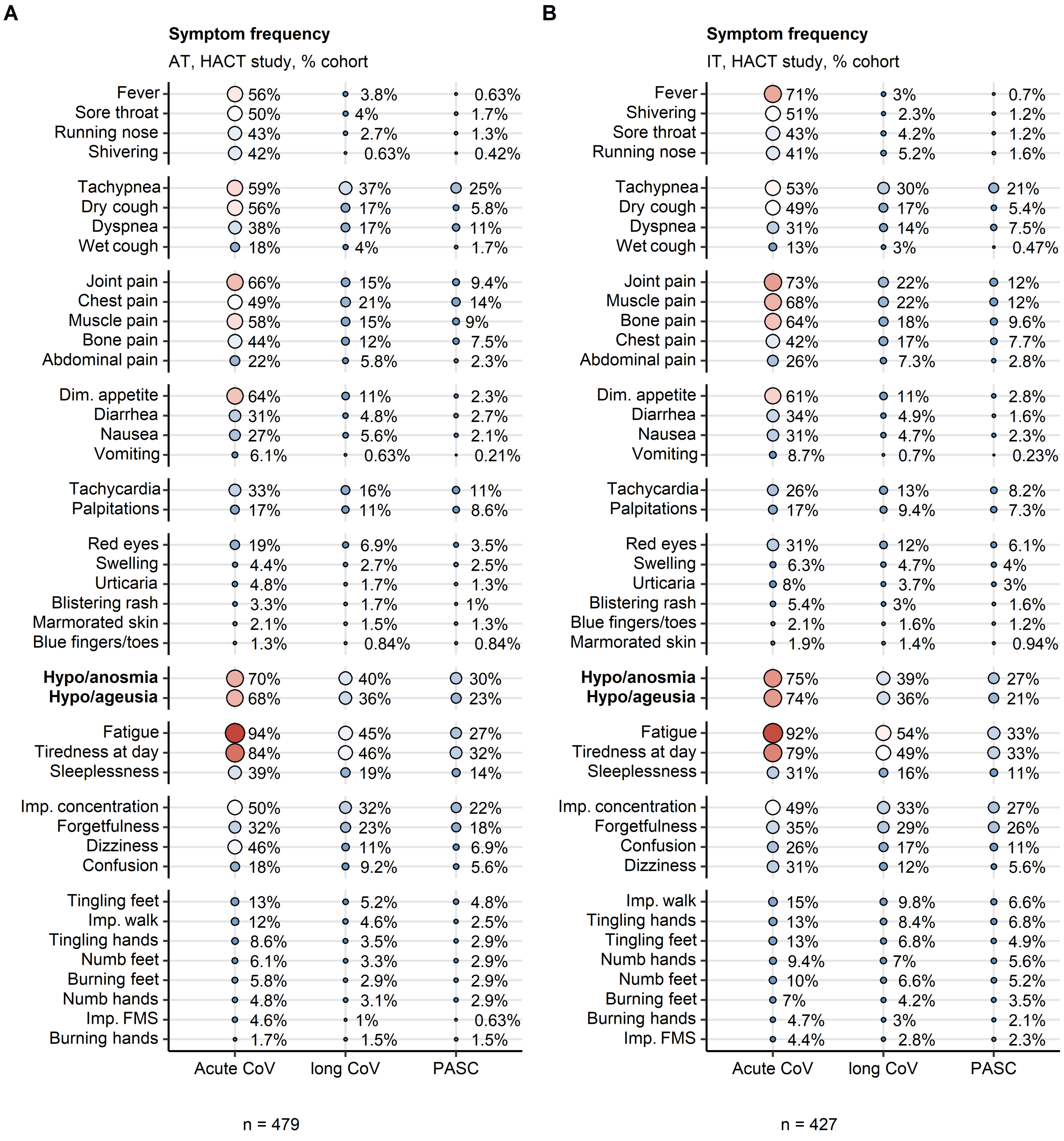


**Supplementary Figure S1. Frequency of COVID-19 symptoms in acute COVID-19, long COVID and PASC in the HACT study.**

Frequency of symptoms of acute COVID-19 (first 14 days post clinical onset), long COVID (≥ 28 days) and PASC (post-acute sequelae of COVID-19, ≥ 90 days) in the AT (Austria, **A**) and IT (Italy, **B**) HACT study cohorts expressed as percentages of the cohort. Point size and color represents the percentage. Numbers of complete observations are indicated under the plot.

Dim. appetite: diminished appetite, Imp. concentration: impaired concentration, Imp. walk: impaired walk, Imp. FMS: impaired fine motor skills.

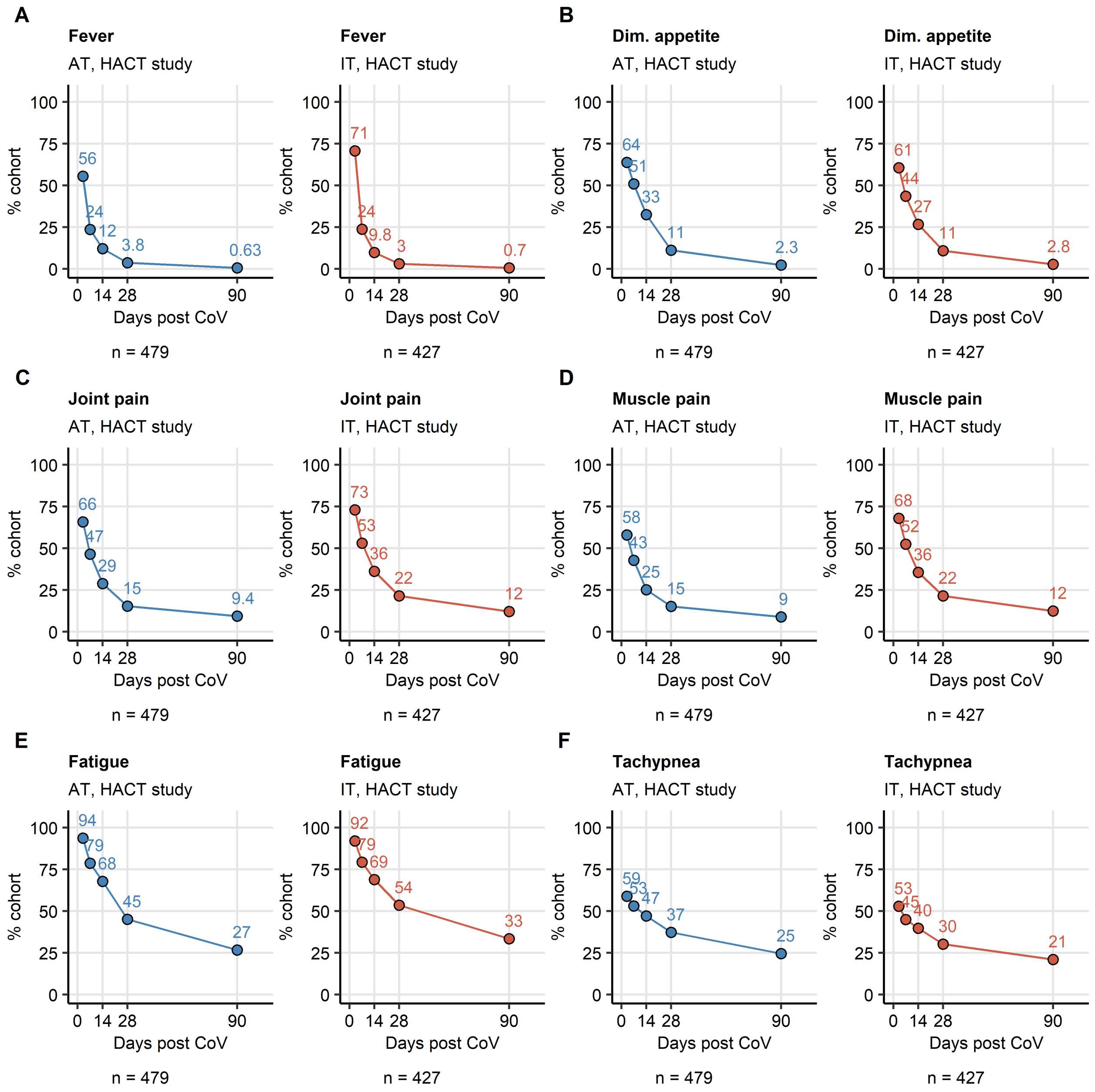


**Supplementary Figure S2. Kinetic of recovery from leading acute COVID-19 symptoms in the HACT study.**

Percentages of individuals with fever (**A**), diminished appetite (**B**), joint pain (**C**), muscle pain (**D**), fatigue (**E**) and tachypnea (**F**) in the AT (Austria) and IT (Italy) HACT study cohorts at particular time points after clinical onset. Numbers of complete observations are indicated under the plots.

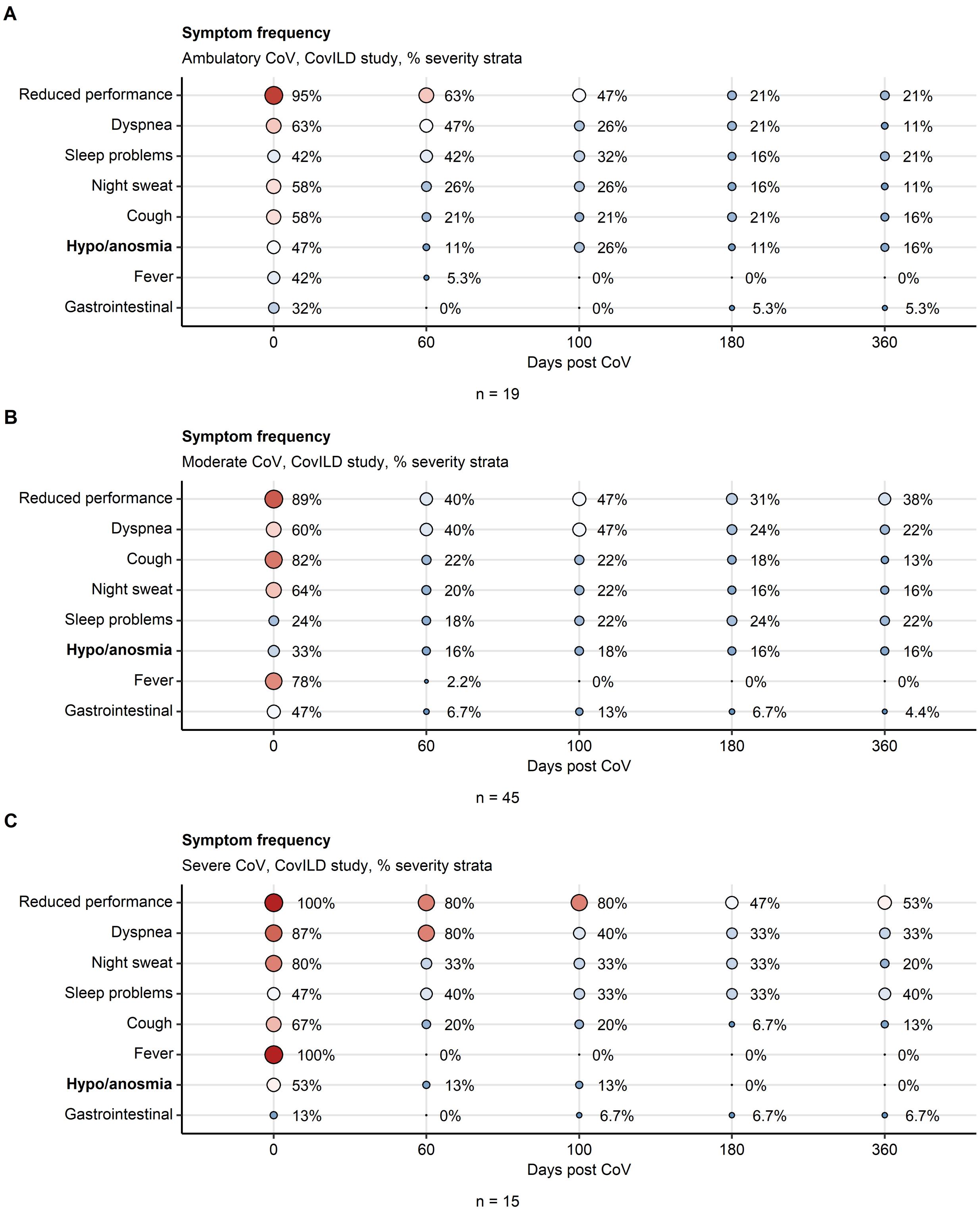
A
Symptom frequency
Ambulatory CoV, CovILD study, % severity strata
Reduced performance 95% 63% 47% 21% 21%
Dyspnea 63% 47% 26% 21% 11%
Sleep problems 42% 42% 32% 16% 21%
Night sweat 58% 26% 26% 16% 11%
Cough 58% 21% 21% 21% 16%
Hypo/anosmia 47% 11% 26% 11% 16%
Fever 42% 5.3% 0% 0% 0%
Gastrointestinal 32% 0% 0% 5.3% 5.3%
0 60 100 180 360
Days post CoV
n = 19
B
Symptom frequency
Moderate CoV, CovILD study, % severity strata
Reduced performance 89% 40% 47% 31% 38%
Dyspnea 60% 40% 47% 24% 22%
Cough 82% 22% 22% 18% 13%
Night sweat 64% 20% 22% 16% 16%
Sleep problems 24% 18% 22% 24% 22%
Hypo/anosmia 33% 16% 18% 16% 16%
Fever 78% 2.2% 0% 0% 0%
Gastrointestinal 47% 6.7% 13% 6.7% 4.4%
0 60 100 180 360
Days post CoV
n = 45
C
Symptom frequency
Severe CoV, CovILD study, % severity strata
Reduced performance 100% 80% 80% 47% 53%
Dyspnea 87% 80% 40% 33% 33%
Night sweat 80% 33% 33% 33% 20%
Sleep problems 47% 40% 33% 33% 40%
Cough 67% 20% 20% 6.7% 13%
Fever 100% 0% 0% 0% 0%
Hypo/anosmia 53% 13% 13% 0% 0%
Gastrointestinal 13% 0% 6.7% 6.7% 6.7%
0 60 100 180 360
Days post CoV
n = 15

**Supplementary Figure S3. Symptom frequency in ambulatory, moderate and severe COVID-19 subsets of the CovILD study.**

Frequency of symptoms during acute COVID-19 and at the 60-, 100-, 180- and 360-day follow-ups in ambulatory (**A**), moderate (**B**) and severe COVID-19 (**C**) participants of the CovILD study expressed as percentages of individuals with the complete longitudinal data set. Point size and color represents the percentage. Numbers of complete observations are indicated under the plots.

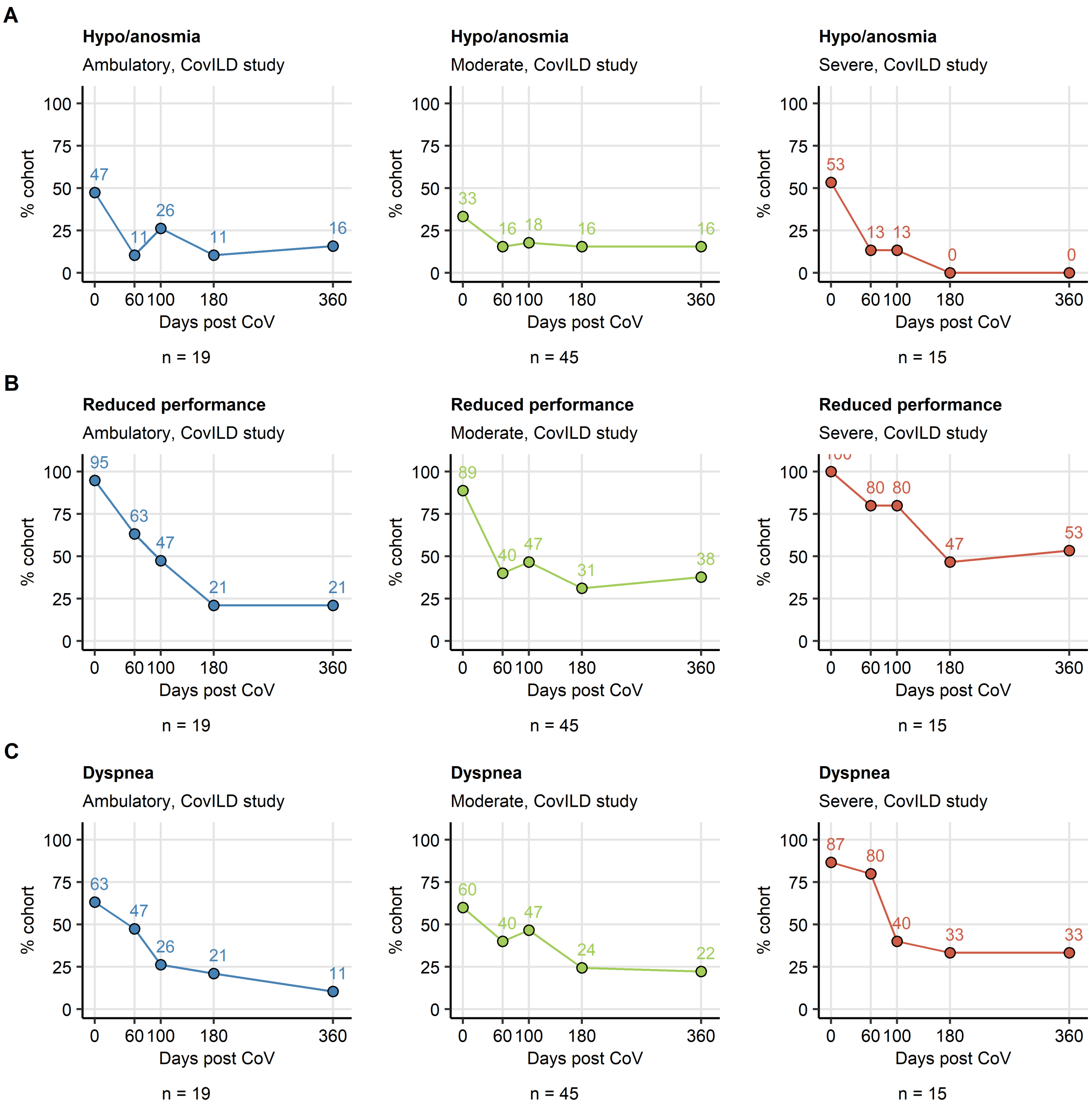


**Supplementary Figure S4. Kinetic of recovery from smell disorders, reduced performance and dyspnea in ambulatory, moderate and severe COVID-19 subsets of the CovILD study.**

Percentages of individuals with the complete longitudinal data set suffering from smell disorders (**A**), reduced physical performance (**B**) and dyspnea (**C**) in the ambulatory, moderate and severe COVID-19 subsets of the CovILD study during acute COVID-19 and at the 60-, 100-, 180- and 360-day follow-ups. Numbers of complete observations are indicated under the plots.

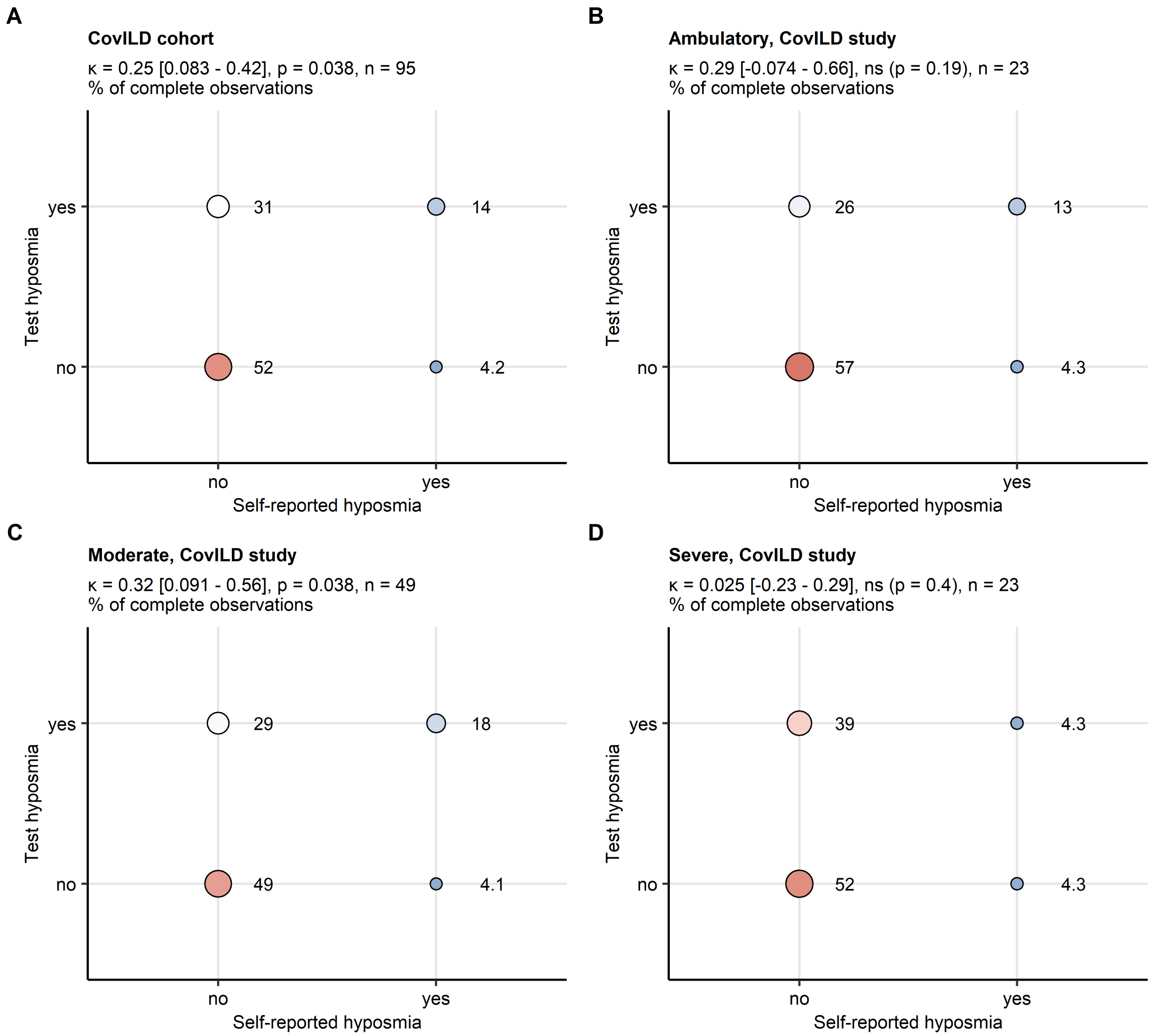


**Supplementary Figure S5. Rates of self-reported hyposmia and hyposmia in sniffing stick test at 3-month post COVID-19 follow-up in the ambulatory, moderate and severe COVID-19 subsets of the CovILD study.**

Association of self-reported and sniffing stick test hyposmia rates was investigated with Cohen's $\kappa$ statistic. Statistical significance was assessed with Wald's Z test corrected for multiple testing with Benjamini-Hochberg method. Percentages of individuals with self-reported and test hyposmia within the entire cohort (**A**), the ambulatory (**B**), moderate (**C**) and severe (**D**) COVID-19 subsets of the CovILD-19 subsets are presented in bubble plots. Point size and color represents the percentage. $\kappa$ values with 95% confidence intervals, p values and numbers of complete observations are indicated in the plot captions.

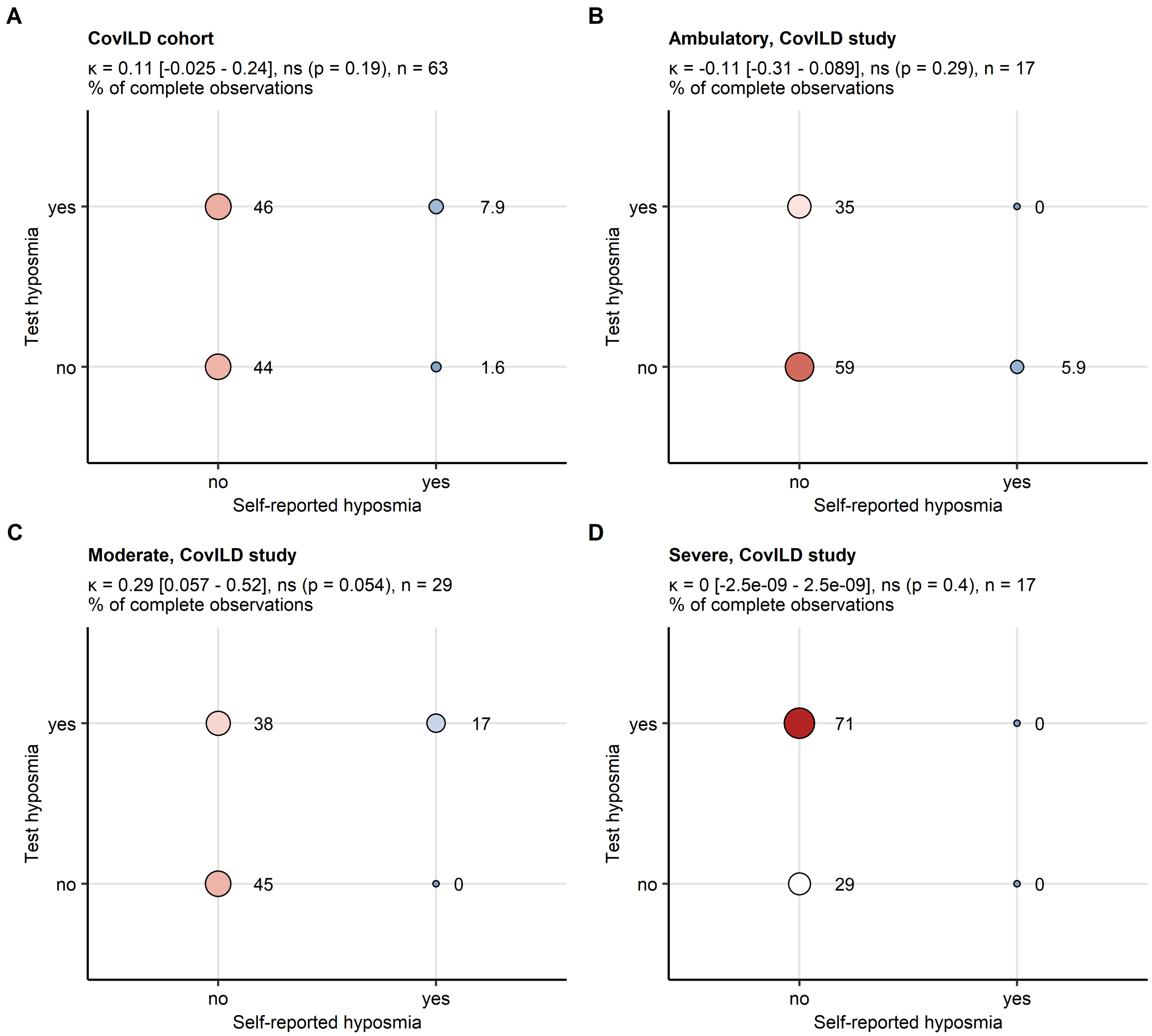


**Supplementary Figure S6. Rates of self-reported hyposmia and hyposmia in sniffing stick test at 1-year post COVID-19 follow-up in the ambulatory, moderate and severe COVID-19 subsets of the CovILD study.**

Association of self-reported and sniffing stick test hyposmia rates was investigated with Cohen's $\kappa$ statistic. Statistical significance was assessed with Wald's Z test corrected for multiple testing with Benjamini-Hochberg method. Percentages of individuals with self-reported and test hyposmia within the entire cohort (**A**), the ambulatory (**B**), moderate (**C**) and severe (**D**) COVID-19 subsets of the CovILD-19 subsets are presented in bubble plots. Point size and color represents the percentage. $\kappa$ values with 95% confidence intervals, p values and numbers of complete observations are indicated in the plot captions.

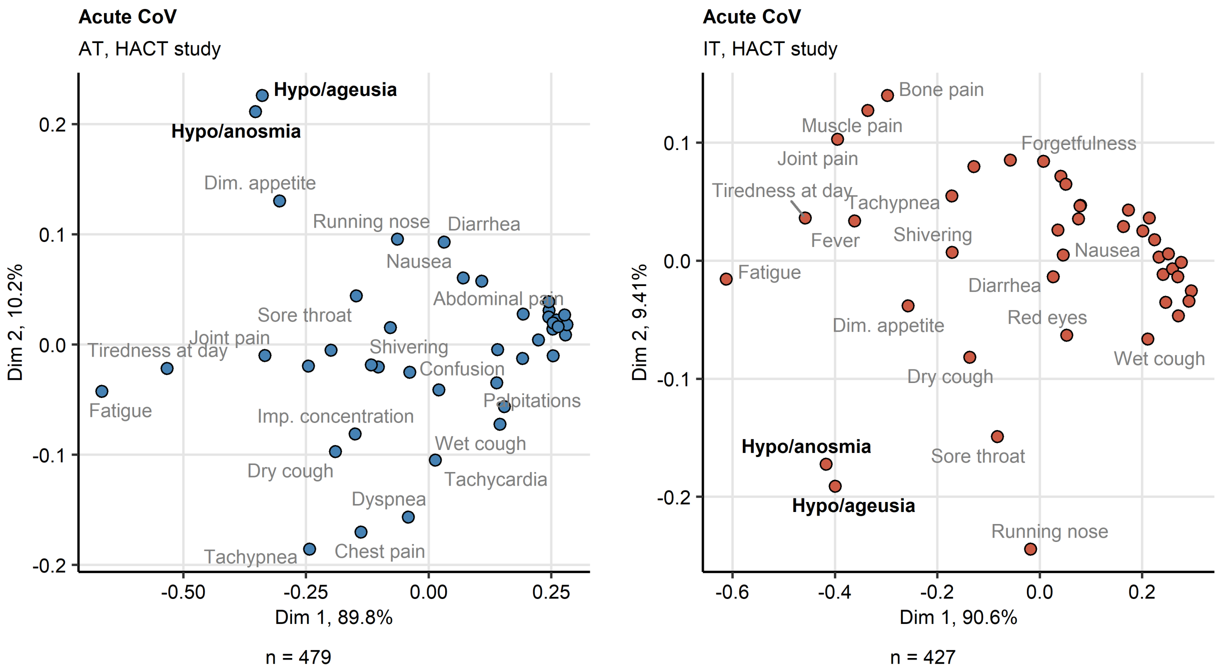


**Supplementary Figure S7. Multi-dimensional scaling analysis of acute COVID-19 symptoms in the HACT study.**

Binary symptom occurrence data for acute COVID-19 (first 14 days after clinical onset) in the HACT study Austria (AT) and Italy (IT) cohorts were subjected to two-dimensional multi-dimensional scaling (MDS) with simple matching distance (SMD) between the symptoms. MDS coordinates are presented in point plots. Selected data points are labeled with the symptom names. Percentages of the data set variance associated with the MDS dimensions are indicated in the plot axes. Numbers of complete observations are indicated under the plots.

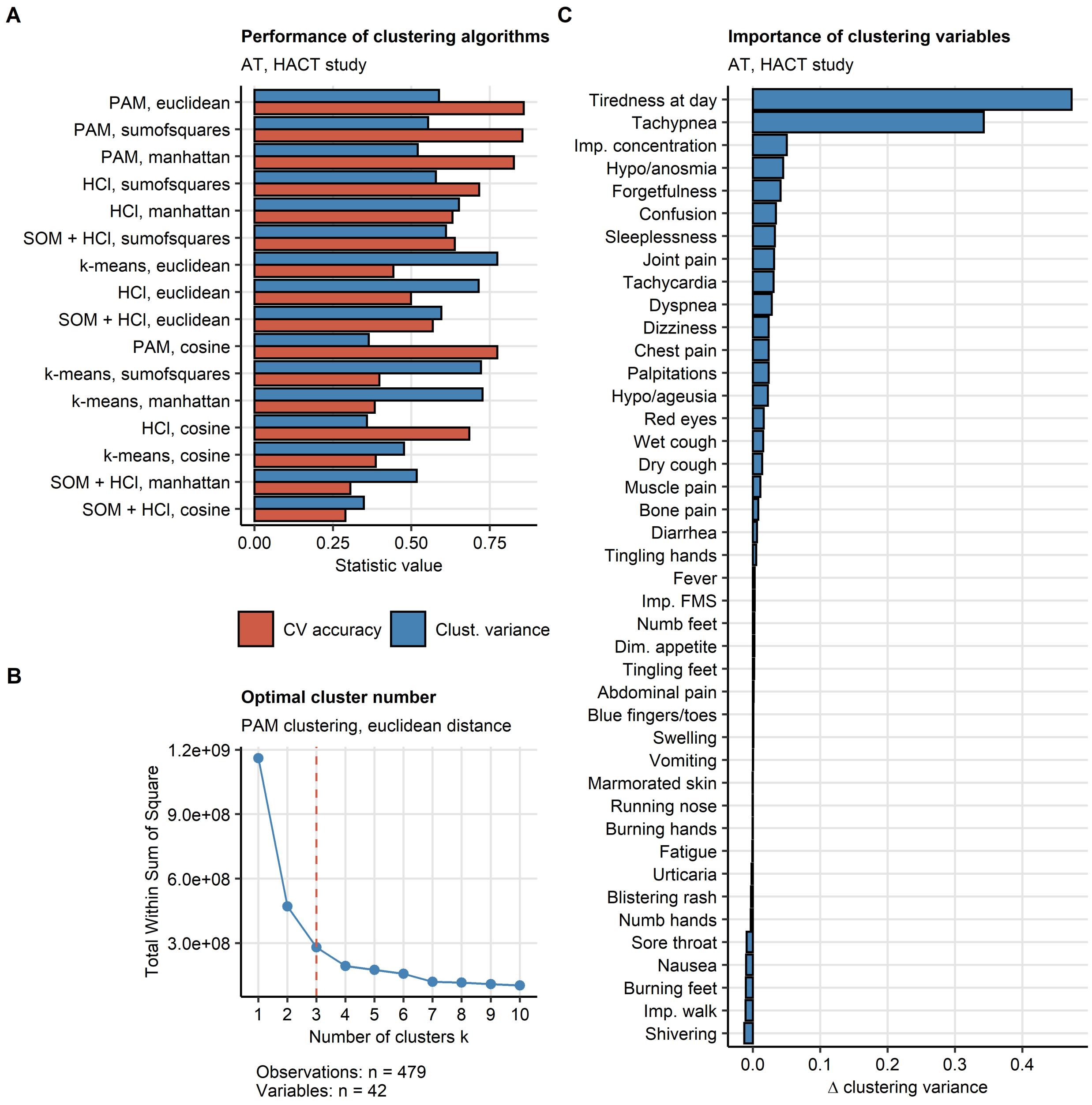


**Supplementary Figure S8. Definition of the COVID-19 recovery clusters and clustering feature importance in the HACT study.**

Individuals of the training Austria (AT) cohort of the HACT study were subjected to clustering in respect to symptom-specific recovery times with the PAM (partitioning around medoids) algorithm and Euclidean distance measure.

**(A)** Comparison of performance of various algorithms (HCl: hierarchical clustering, SOM + HCl: combined self-organizing map and hierarchical clustering, k-means) and distance statistic in clustering of the training data set investigated by clustering variance (ratio of total between-cluster sum of squares to total sum of squares) and cluster assignment accuracy in 10-fold cross-validation (CV).

**(B)** Determination of the optimal cluster number in the PAM clustering of the training cohort by the bend of the total within-cluster sum of squares curve.

**(C)** Permutation importance of the clustering features (symptoms) for clustering of the training cohort expressed as the difference in clustering variance (ratio of total between-cluster sum of squares to total sum of squares) between the initial clustering object and the clustering object with the given variable reshuffled at random.

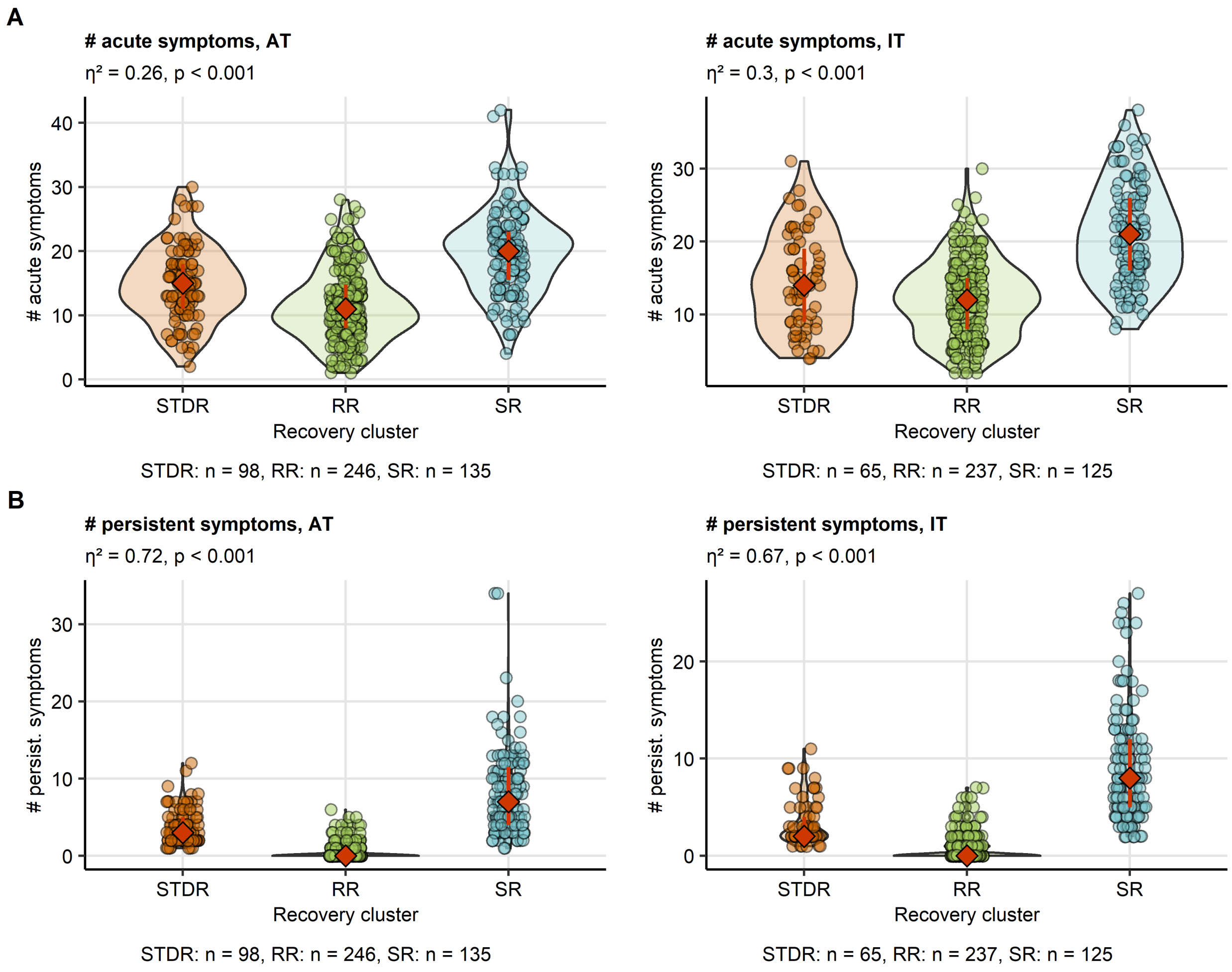


**Supplementary Figure S9. Numbers of acute COVID-19 and long COVID symptoms in the HACT study recovery clusters.**

The recovery clusters were defined in the training Austria cohort (AT, **Supplementary Figure S8**) and the cluster assignment predicted in the test Italy cohort (IT, **Figure 5**). Differences in numbers of acute COVID-19 (first 14 days after clinical onset, **A**) and long COVID symptoms (≥ 28 days, **B**) between the clusters were assessed by Kruskal-Wallis test and $\eta^2$ effect size statistic. P values were corrected for multiple testing with Benjamini-Hochberg method. Symptom counts are presented in violin plots. Points represent single observations, orange diamonds with whiskers code for medians and interquartile ranges. Effect size statistics and p values are indicated in the plot caption. Numbers of complete observations are displayed under the plots.

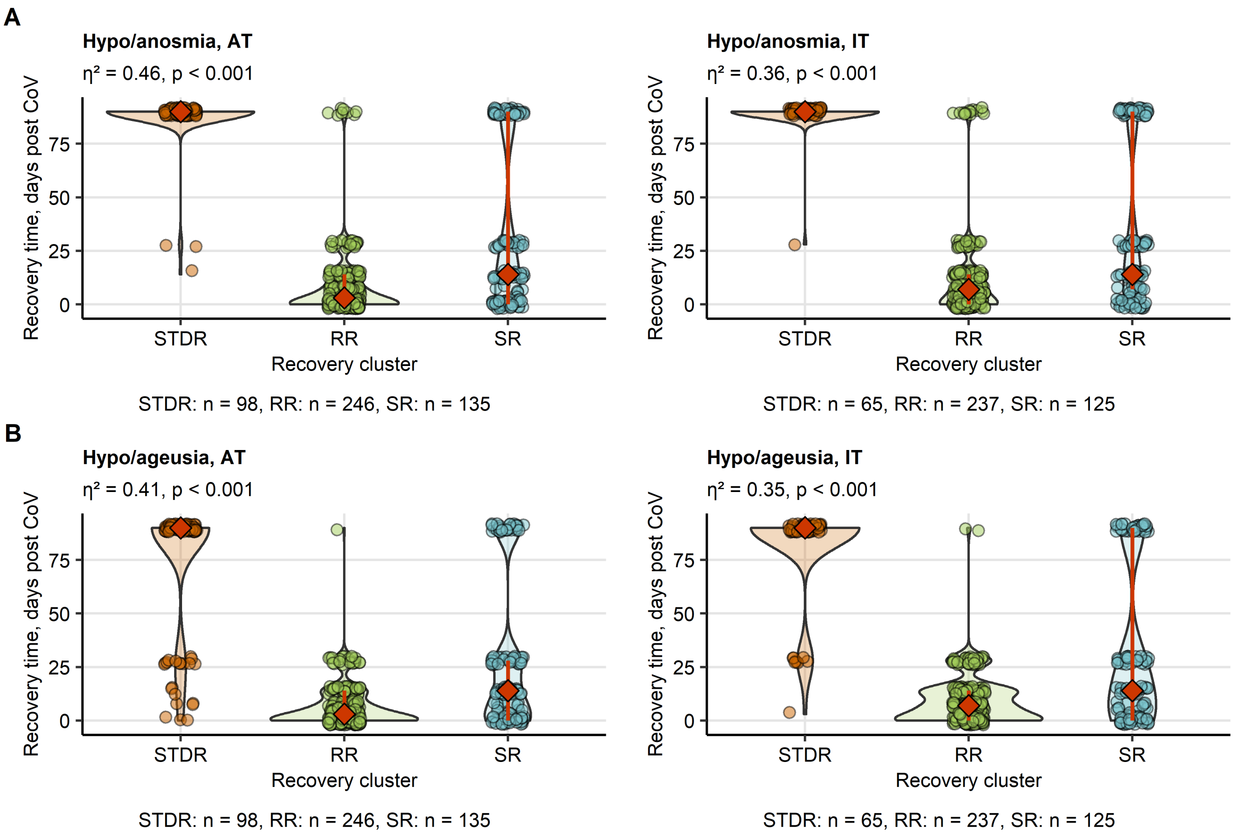


**Supplementary Figure S10. Recovery from smell and taste disorders in the HACT study recovery clusters**

The recovery clusters were defined in the training Austria cohort (AT, **Supplementary Figure S8**) and the cluster assignment predicted in the test Italy cohort (IT, **Figure 5**). Differences in recovery times for smell (**A**) and taste disorders (**D**) between the clusters were assessed by Kruskal-Wallis test and $\eta^2$ effect size statistic. P values were corrected for multiple testing with Benjamini-Hochberg method. Symptom counts are presented in violin plots. Points represent single observations, orange diamonds with whiskers code for medians and interquartile ranges. Effect size statistics and p values are indicated in the plot caption. Numbers of complete observations are displayed under the plots.

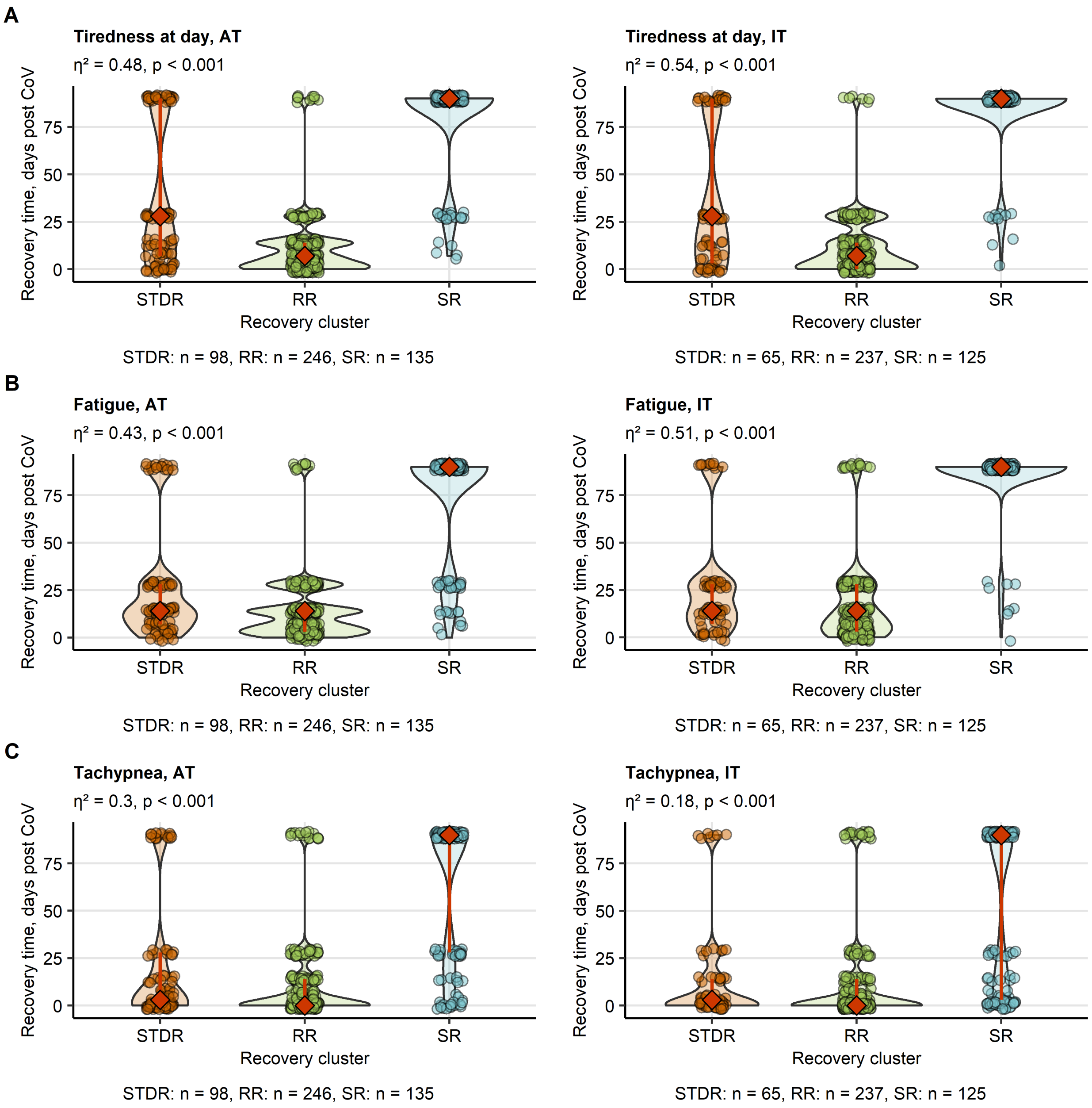


**Supplementary Figure S11. Recovery from fatigue, tiredness and tachypnea in the HACT study recovery clusters.**

The recovery clusters were defined in the training Austria cohort (AT, **Supplementary Figure S8**) and the cluster assignment predicted in the test Italy cohort (IT, **Figure 5**). Differences in recovery times for fatigue (**A**), day tiredness (**B**) and tachypnea (**C**) between the clusters were assessed by Kruskal-Wallis test and $\eta^2$ effect size statistic. P values were corrected for multiple testing with Benjamini-Hochberg method. Symptom counts are presented in violin plots. Points represent single observations, orange diamonds with whiskers code for medians and interquartile ranges. Effect size statistics and p values are indicated in the plot caption. Numbers of complete observations are displayed under the plots.

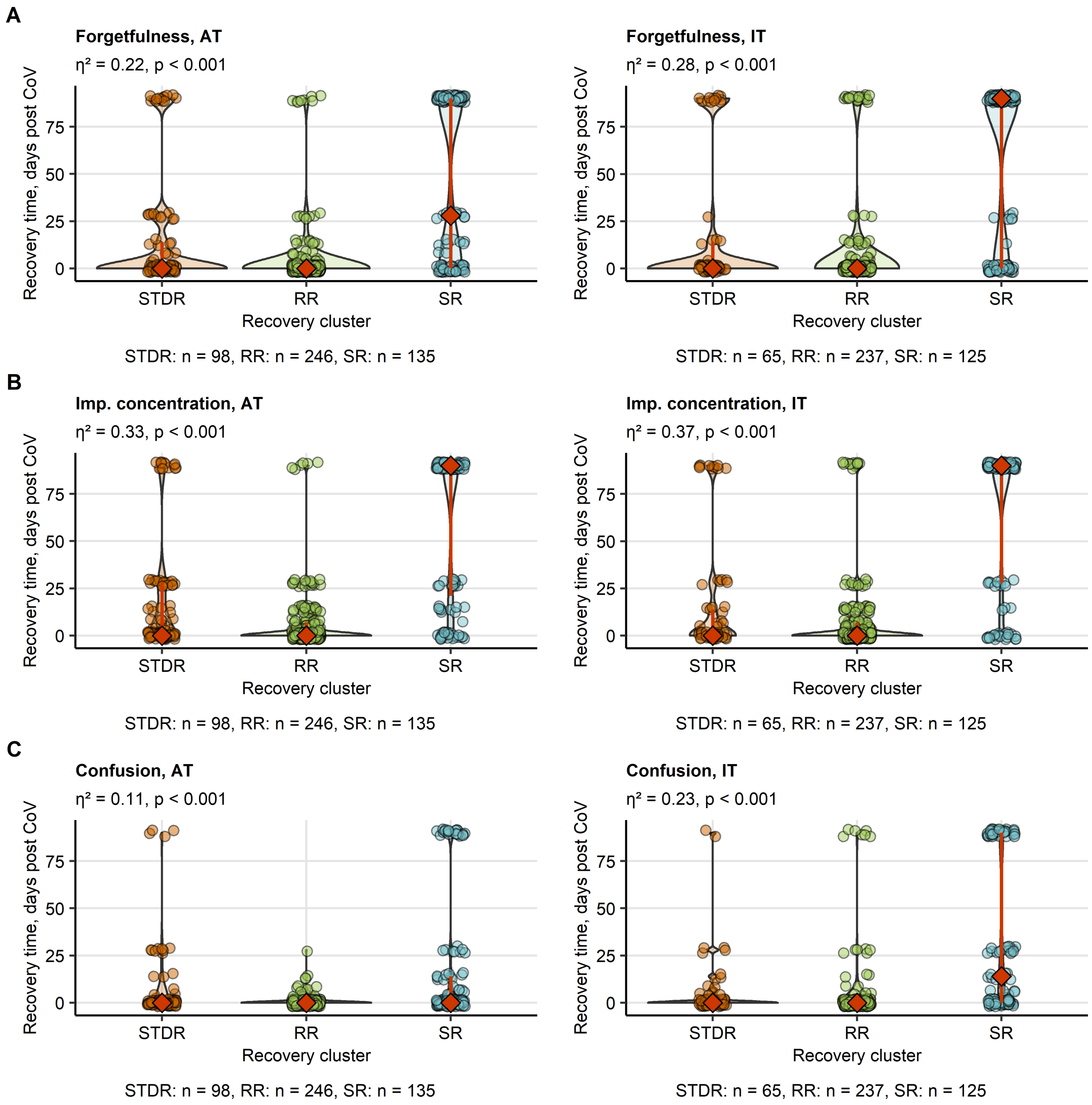


**Supplementary Figure S12. Recovery from neurocognitive symptoms of COVID-19 in the HACT study recovery clusters.**

The recovery clusters were defined in the training Austria cohort (AT, **Supplementary Figure S8**) and the cluster assignment predicted in the test Italy cohort (IT, **Figure 5**). Differences in recovery times for forgetfulness (**A**), concentration impairment (**B**) and confusion (**C**) between the clusters were assessed by Kruskal-Wallis test and $\eta^2$ effect size statistic. P values were corrected for multiple testing with Benjamini-Hochberg method. Symptom counts are presented in violin plots. Points represent single observations, orange diamonds with whiskers code for medians and interquartile ranges. Effect size statistics and p values are indicated in the plot caption. Numbers of complete observations are displayed under the plots.

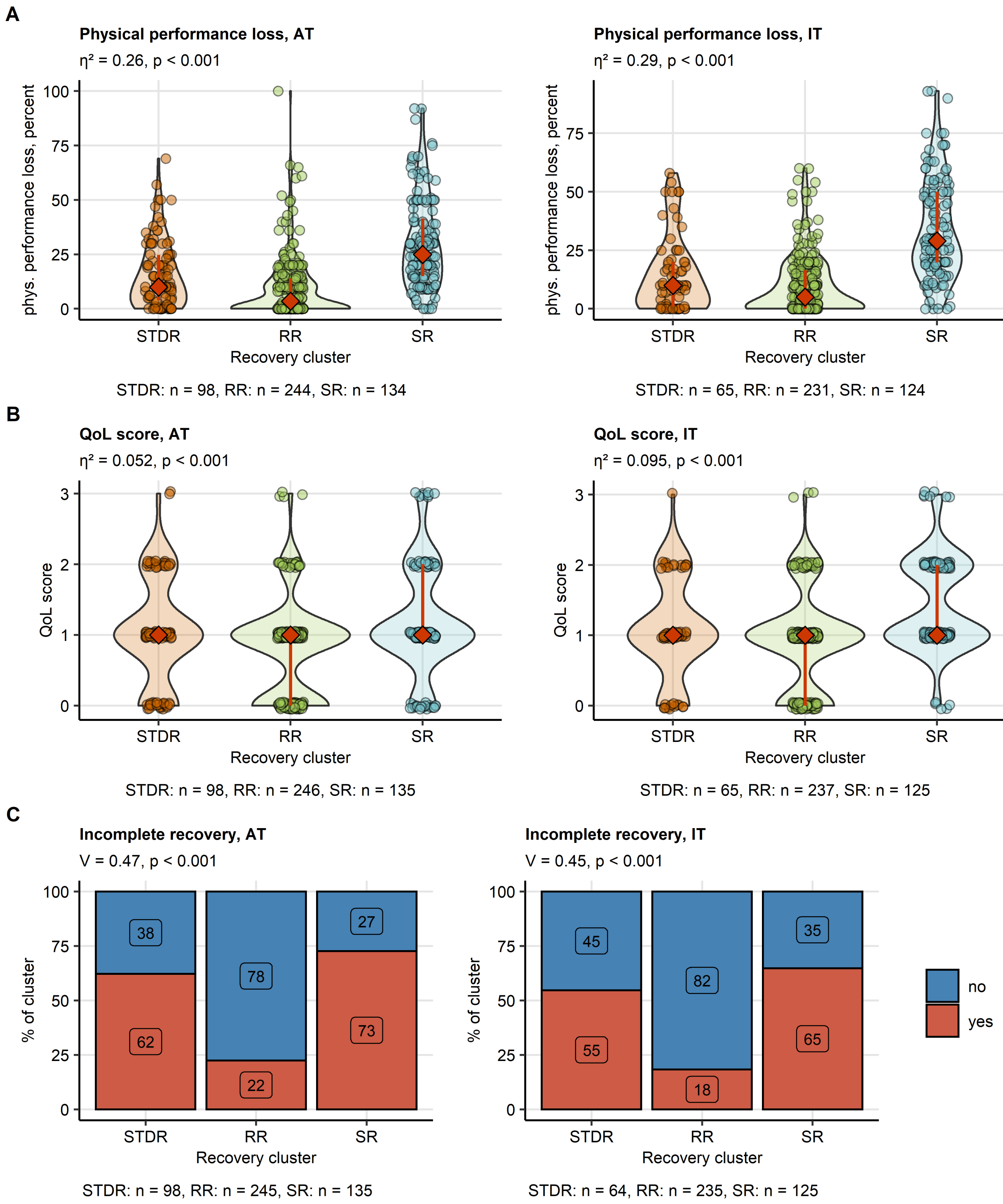


**Supplementary Figure S13. Physical performance loss, impairment of quality of life and self-perceived complete convalescence at the survey completion in the HACT study recovery clusters.**

The recovery clusters were defined in the training Austria cohort (AT, **Supplementary Figure S8**) and the cluster assignment predicted in the test Italy cohort (IT, **Figure 5**). Differences in self-reported physical performance loss (**A**) and scoring of impairment of quality of life (QoL, **B**) at the survey completion between the clusters were assessed by Kruskal-Wallis test and $\eta^2$ effect size statistic. The scoring data were presented in violin plots. Points represent single observations, orange diamonds with whiskers code for medians and interquartile ranges. Differences in frequency of self-perceived complete convalescence (**C**) between the clusters were investigated by $\chi^2$ test and Cramer V effect size statistic. The frequencies are presented as bar plots. P values were corrected for multiple testing with Benjamini-Hochberg method. Effect size statistics and p values are indicated in the plot caption. Numbers of complete observations are displayed under the plots.

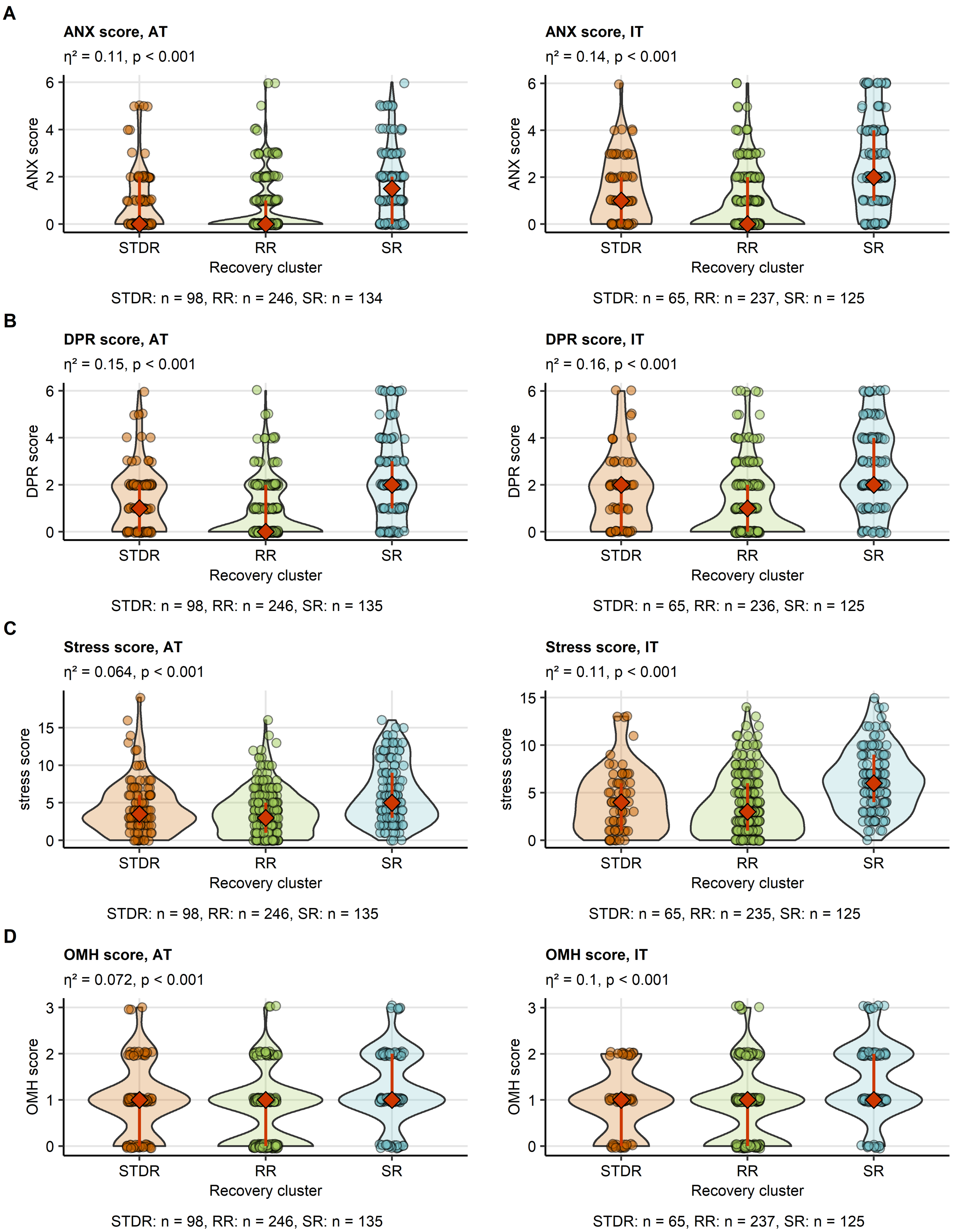
A
ANX score, AT
η² = 0.11, p < 0.001
ANX score
Recovery cluster
STDR
RR
SR
STDR: n = 98, RR: n = 246, SR: n = 134
ANX score, IT
η² = 0.14, p < 0.001
STDR: n = 65, RR: n = 237, SR: n = 125
B
DPR score, AT
η² = 0.15, p < 0.001
DPR score
STDR: n = 98, RR: n = 246, SR: n = 135
DPR score, IT
η² = 0.16, p < 0.001
STDR: n = 65, RR: n = 236, SR: n = 125
C
Stress score, AT
η² = 0.064, p < 0.001
stress score
STDR: n = 98, RR: n = 246, SR: n = 135
Stress score, IT
η² = 0.11, p < 0.001
STDR: n = 65, RR: n = 235, SR: n = 125
D
OMH score, AT
η² = 0.072, p < 0.001
OMH score
STDR: n = 98, RR: n = 246, SR: n = 135
OMH score, IT
η² = 0.1, p < 0.001
STDR: n = 65, RR: n = 237, SR: n = 125

**Supplementary Figure S14. Rating of anxiety, depression, overall mental health impairment and stress at the survey completion in the HACT study recovery clusters.**

The recovery clusters were defined in the training Austria cohort (AT, **Supplementary Figure S8**) and the cluster assignment predicted in the test Italy cohort (IT, **Figure 5**). Differences in rating of anxiety (ANX, **A**), depression (DPR, **B**), stress (**C**) and self-reported overall mental health impairment (OMH, **D**) at the survey completion between the clusters were assessed by Kruskal-Wallis test and $\eta^2$ effect size statistic. P values were corrected for multiple testing with Benjamini-Hochberg method. The scoring data were presented in violin plots. Points represent single observations, orange diamonds with whiskers code for medians and interquartile ranges. Effect size statistics and p values are indicated in the plot caption. Numbers of complete observations are displayed under the plots.
